# Recruitment of highly functional SARS-CoV-2-specific CD8^+^ T cell receptors mediating cytotoxicity of virus-infected target cells in non-severe COVID-19

**DOI:** 10.1101/2021.07.20.21260845

**Authors:** Karolin I. Wagner, Laura M. Mateyka, Sebastian Jarosch, Vincent Grass, Simone Weber, Kilian Schober, Monika Hammel, Teresa Burrell, Behnam Kalali, Holger Poppert, Henriette Beyer, Sophia Schambeck, Stefan Holdenrieder, Andrea Strötges-Achatz, Verena Haselmann, Michael Neumaier, Johanna Erber, Alina Priller, Sarah Yazici, Hedwig Roggendorf, Marcus Odendahl, Torsten Tonn, Andrea Dick, Klaus Witter, Hrvoje Mijočević, Ulrike Protzer, Percy A. Knolle, Andreas Pichlmair, Claudia S. Crowell, Markus Gerhard, Elvira D’Ippolito, Dirk H. Busch

**Affiliations:** Institute for Medical Microbiology, Immunology and Hygiene, Technical University of Munich (TUM), Munich, Germany; Institute of Virology, School of Medicine, Technical University of Munich (TUM) and Helmholtz Zentrum Muenchen, Munich, Germany; Mikrobiologisches Institut – Klinische Mikrobiologie, Immunologie und Hygiene, Universitätsklinikum Erlangen, Friedrich-Alexander-Universität (FAU) Erlangen-Nürnberg, Germany; Department of Neurology, Helios Klinikum München West, Munich, Germany; Institute of Laboratory Medicine, Munich Biomarker Research Center, Deutsches Herzzentrum München, Technical University of Munich (TUM), Munich, Germany; Department of Clinical Chemistry, University Medicine Mannheim, Medical Faculty Mannheim of the University of Heidelberg, Mannheim, Germany; Department of Internal Medicine II, University Hospital rechts der Isar, Technical University of Munich (TUM), Munich, Germany; Institute of Molecular Immunology and Experimental Oncology, School of Medicine, Technical University of Munich (TUM), Munich, Germany; Experimental Transfusion Medicine, Medical Faculty Carl Gustav Carus, TU Dresden, Germany; Institute for Transfusion Medicine Dresden, German Red Cross Blood Donation Service North-East, Dresden, Germany; Laboratory of Immunogenetics and Molecular Diagnostics, Department of Transfusion Medicine, Cell Therapeutic Agents and Hemostaseology, LMU Munich, Munich, Germany; German Center for Infection Research (DZIF), partner site Munich, Munich, Germany

## Abstract

T cell immunity is crucial for the control of severe acute respiratory syndrome coronavirus 2 (SARS-CoV-2) infections and has been widely characterized on a quantitative level. In contrast, the quality of such T cell responses has been poorly investigated, in particular in the case of CD8^+^ T cells. Here, we explored the quality of SARS-CoV-2-specific CD8^+^ T cell responses in individuals who recovered from mild symptomatic infections, through which protective immunity should develop, by functional characterization of their T cell receptor (TCR) repertoire. CD8^+^ T cell responses specific for SARS-CoV-2-derived epitopes were low in frequency but could be detected robustly early as well as late - up to twelve months - after infection. A pool of immunodominant epitopes, which accurately identified previous SARS-CoV-2 infections, was used to isolate TCRs specific for epitopes restricted by common HLA class I molecules. TCR-engineered T cells showed heterogeneous functional avidity and cytotoxicity towards virus-infected target cells. High TCR functionality correlated with gene signatures of T cell function and activation that, remarkably, could be retrieved for each epitope:HLA combination and patient analyzed. Overall, our data demonstrate that highly functional HLA class I TCRs are recruited and maintained upon mild SARS-CoV-2 infection. Such validated epitopes and TCRs could become valuable tools for the development of diagnostic tests determining the quality of SARS-CoV-2-specific CD8^+^ T cell immunity, and thereby investigating correlates of protection, as well as to restore functional immunity through therapeutic transfer of TCR-engineered T cells.

## INTRODUCTION

Severe acute respiratory syndrome coronavirus 2 (SARS-CoV-2) is a new beta coronavirus responsible for the Coronavirus Disease 2019 (COVID-19), a pathological condition that can progress to severe pneumonia and a fatal outcome^1,2^. The adaptive immune system plays a critical role in SARS-CoV-2 infections. In most cases, T and B cells react quickly and in a coordinated manner to the infection^3–5^ and develop a robust memory pool detectable months after exposure regardless of disease severity^6–8^. In line with that, individuals who recovered from COVID-19 experienced low reinfection rates, in particular for symptomatic infections^9–11^. In contrast, severe clinical manifestations occurred in the less frequent scenario where immune responses were suboptimal – i.e. in elderly individuals where antigen presentation is less efficient and the pool of naïve T cells is scarce^4,12^. Despite the increasing understanding of SARS-CoV-2 immunity, a clear correlate of protection is still missing. This is of paramount importance as it would allow for the identification of individuals with minimal risk of reinfection as well as the minority of patients with high risk of developing severe symptoms due to lack of adequate immunity. In addition, it would provide relevant platforms or tools for the validation of candidate vaccines, including immunity towards virus variants.

Antibody titers have been extensively used to describe SARS-CoV-2 adaptive immunity as their detection is suitable for high-throughput testing and intrinsically mirrors an adequate recruitment of CD4^+^ T cells. However, the role of antibodies in protective immunity is still controversial. Seropositive convalescent individuals showed lower risk of reinfections^11,13,14^ but neither total nor neutralizing antibody titers protected from severe symptoms during primary infections^4,15^. In addition, the resolution of SARS-CoV-2 infections in individuals unable to produce antibodies^16^ further indicated that other immune compartments are rather essential for protective immunity.

Early induction of SARS-CoV-2-specific T cells was shown to associate with milder diseases and less prolonged virus shedding^4,17^ and, remarkably, depletion of CD8^+^ T cells abrogated protectiveness against re-challenge in pre-clinical models^18^. Altogether, this evidence strongly supports a key role of T cells in the control and resolution of SARS-CoV-2 infections. This should hold true in particular for CD8^+^ T cells due to their unique contribution in protecting from intracellular pathogens^19^ by the direct killing of target cells. Of similar importance, T cells persist longer than waning antibodies^8,20^, thus being more informative about the long-term maintenance of functional SARS-CoV-2 immunity.

SARS-CoV-2-specific T cell immunity has been widely characterized quantitatively^6,8,12,21,22^. However, little is known about their quality, and thereby determinants of protection. This lack of knowledge in the COVID-19 field mainly derives from the broad use of single dose peptide mixes (15-mers) for T cell analyses, which are often preferred as they can be designed easily to cover entire open reading frames (ORFs). Their usage was highly valuable to gain information on the magnitude and breadth of SARS-CoV-2 T cell responses in a remarkably short time^8,22^ but at the expense of precision, i.e. specificity and quality of single-epitope responses. Indeed, the epitopes responsible for the observed T cell reactivity are often unknown and peptide mixes are usually used at a single high concentration, which hinders discrimination of high (protective) and low (suboptimal) functional T cells. This information has become extremely relevant as i) high frequency of cross-reactivity with common cold coronaviruses was reported, even though mainly for CD4^+^ T cells^23–27^, and ii) suboptimal - but high-frequency - T cell responses were observed in severe COVID-19 patients due to recruitment of low functional cross-reactive memory T cells rather than highly specific naïve T cells^12^. Finally, CD8^+^ T cell responses are still under-represented, and thereby less investigated, as 15-mer peptides primarily stimulate CD4^+^ T cells^25^. Altogether, this evidence underlines how the detection and magnitude of T cell responses is not always equivalent to functionality.

Thus, we decided to investigate in depth the quality of SARS-CoV-2-specific CD8^+^ T cells through the analyses of their T cell receptor (TCR) repertoire. A TCR is the fingerprint of a T cell and determines its specificity, functionality and fate. Furthermore, preclinical studies have shown that highly functional TCRs drive the establishment of protective immunity in infectious diseases, as they dominate primary infections^28^ and react faster to recall infections^29^. By using SARS-CoV-2 immunodominant epitopes restricted to the most common Human Leucocyte Antigen (HLA) class I molecules, we accessed the SARS-CoV-2 epitope-specific TCR repertoire in convalescent individuals who experienced mild symptoms and for whom, thereby, a protective immunity should have established. We identified highly functional TCRs that, when engineered in primary T cells, conferred high sensitivity to epitope stimulation and cytolytic capacity toward target cells which were infected with viable SARS-CoV-2 virus. Interestingly, TCR functionality correlated with a gene signature of recent activation that we could retrieve from each epitope:HLA combination analyzed. Overall, we show that highly functional, and thereby protective, TCRs are recruited in CD8^+^ T cell responses against SARS-CoV-2 during non-severe disease.

## RESULTS

### Detection of SARS-CoV-2-specific CD8^+^ T cells through 9-mer peptide pool stimulation

To study SARS-CoV-2-specific CD8^+^ T cell responses, we designed a pool of SARS-CoV-2 peptides (9-mer) predicted to be immunogenic for the most common HLA class I molecules (HLA-A*01:01, A*02:01, A*03:01, A*11:01, A*24:02, B*07:02, B*08:01, B*35:01). This HLA combination covers 73% of the European Caucasian population (cumulative HLA allele frequency, allelefrequencies.net, similar to^30^). Briefly, we performed *in silico* predictions for peptides of 8-11 amino acid length derived from all ORFs of the SARS-CoV-2 genome and analyzed for their MHC Class I binding affinity to the selected set of HLAs. Peptides with high prediction for stable HLA binding were further screened for additional parameters (i.e. proteasomal cleavage, immunogenicity and transport to the cell surface), which are important prerequisites for a proper antigen surface presentation. We finally selected 40 candidates, among which nine showed 100% homology with SARS-CoV and 31 were unique for SARS-CoV-2 (Supplementary Table 1). No homology was found with published epitopes (IEBD databank) from “common cold” coronaviruses (229E, NL63, HKU1, OC43, degree of homology higher than 70%).

SARS-CoV-2 T cell responses contract and become barely detectable *ex vivo* within a few weeks after infection^31^. Our cohort of convalescent individuals (PCR+ with mild course of the disease and no need of hospitalization, referred to as mild COVID-19 from now on) comprised of blood sampled at least 30-50 days after infection. Therefore, we tested the sensitivity of *ex vivo* responses to a commercially available peptide mix of the Spike (S) protein (Peptivator S, 15-mer mix) and, as expected, we detected low frequency but reliable T cell responses in most mild COVID-19, despite mainly in CD8^-^ T cells (Supplementary Fig. 1). Stimulation with our 9-mer pool, however, indicated detectable T cell responses in few individuals where the size of frequencies and robustness of detection were in addition suboptimal (Supplementary Fig. 1), primarily due to the small size of our pool (4-fold lower number of peptides than Peptivator S pool). Thus, in order to detect such extremely low frequency SARS-CoV-2 reactive CD8^+^ T cell populations, we adapted an expansion protocol where T cells are stimulated with autologous pulsed PBMCs and *in vitro* expanded prior re-challenge and T cell functional analyses^32^ (Fig. 1A). We successfully observed SARS-CoV-2 T cell responses after expansion within a set of mild COVID-19 subjects; furthermore, as expected by the design of the 9-mer peptide pool, primarily CD8^+^ but not CD8^-^ T cells responded to the stimulation (Supplementary Fig. 2).

**Figure 1.**
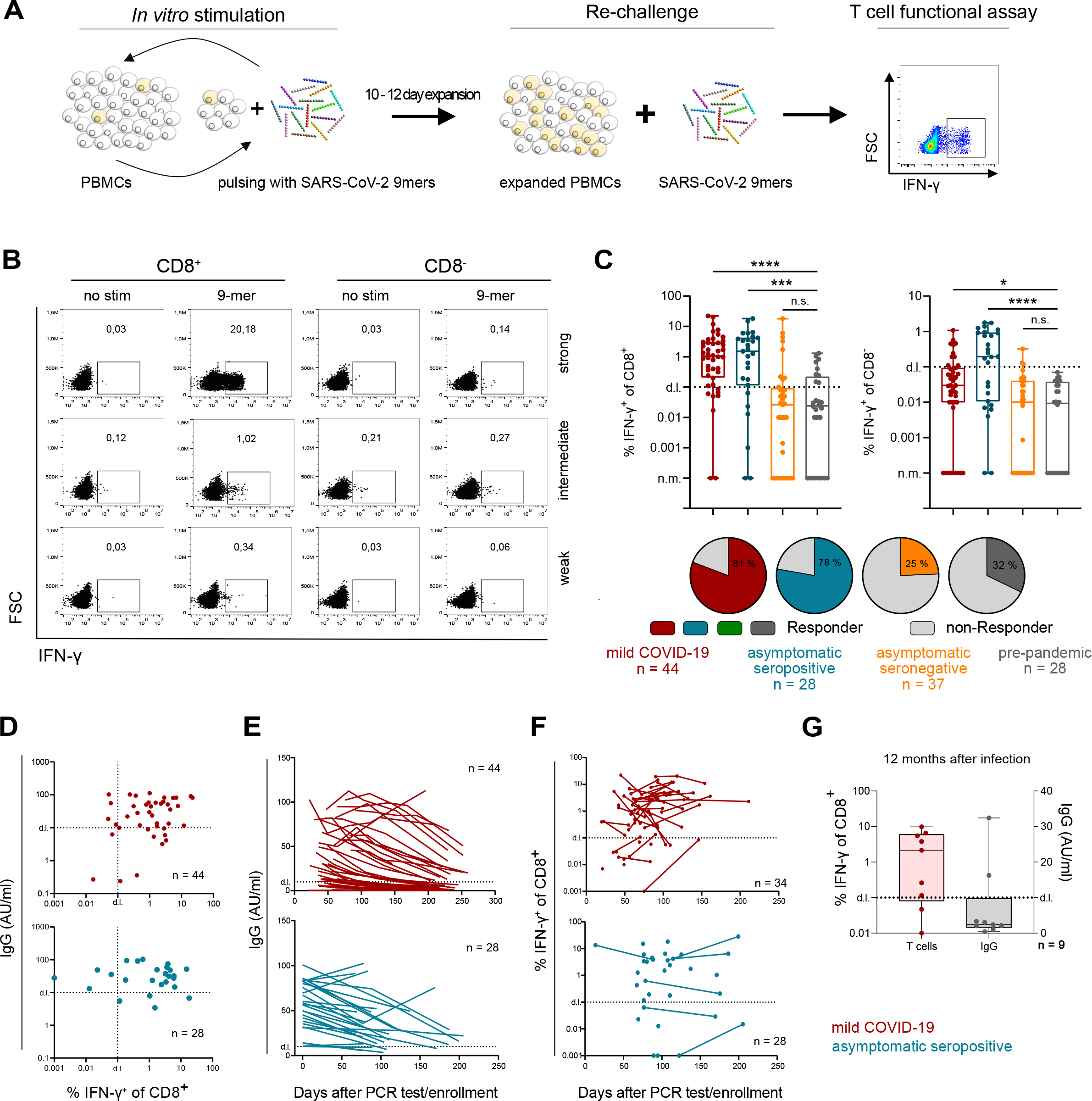
Long term-persistence of SARS-CoV-2-specific CD8^+^ T cells in mild COVID-19 and asymptomatic seropositive individuals. **A)** Schematic overview of the *in vitro* expansion protocol for detection of CD8^+^ T cell responses. 20 % of PBMCs were pulsed with 10 µg/µl SARS-CoV-2 9-mer peptide pool and co-cultured with the remaining 80 % of PBMCs for 10-12 days. 50 IU/ml IL-2 was added every 3-4 days. After expansion, PBMCs were re-challenged with 1 µg/ml crude 9-mer peptide pool for 4 h, and T cell reactivity measured via quantification of IFN-γ release. **B-C)** 5 × 10^6^ PBMCs were treated as described in A. Depicted are representative raw data (B) and quantification (C) of IFN-γ-releasing T cells upon 9-mer peptide pool re-stimulation post-expansion. Responder were identified respective to the non-stimulated negative control and background subtraction. Statistical analyses were performed via one-way Anova with Kurskall-Wallis test (* p value < 0.05, **** p value< 0.001). **D-G)** SARS-CoV-2-specific IgG and CD8^+^ T cells were measured for mild COVID-19 (red) and asymptomatic seropositive (blue) individuals at their first follow-up visit (62,4± 20,4 days after PCR confirmed infection and 102,6± 24,5 days after study enrollment, respectively) (D), throughout the observation (connected lines indicates different time points for the same individual) (E-F) and after one year from PCR confirmed infection (G). For flow cytometry analyses, reactive cells were pre-gated on CD3^+^ living lymphocytes.

### SARS-CoV-2-specific CD8^+^ T cell responses persists long-term in convalescent and asymptomatic seropositive individuals

After validating the robustness of our 9-mer pool in stimulating SARS-CoV-2-specific CD8^+^ T cell responses, we next studied such T cell responses across four distinct cohorts: 53 mild COVID-19; 28 asymptomatic seropositive individuals; 37 asymptomatic individuals who continuously tested seronegative for SARS-CoV-2 IgG antibodies throughout the observation period; 28 unexposed individuals for whom blood was collected before the outbreak (Table 2). Mild COVID-19 and asymptomatic seropositive individuals showed strong, although variable, CD8^+^ T cell responses with 81% and 78% response rate, respectively (detection limit set to 0.1% IFN-γ^+^ CD8^+^/CD8^-^ T cells after background subtraction). In contrast, CD8^+^ T cell responses were observed in a small proportion of asymptomatic seronegative and unexposed individuals, with very weak responses especially for pre-pandemic donors (Fig. 1 B-C). T cell responses could develop in absence of antibody production^6^, supporting our findings in the asympomatic seronegative individuals. Reactivity in pre-pandemic individuals, instead, has been explained as cross-reactive T cells^26,27,33^ or associated to an exceptionally high naïve precursor frequency^34^. As before, CD8^+^ T cells dominated the overall T cell responses after *in vitro* peptide expansion (Fig. 1 C and Supplementary Fig. 3). In order to exclude a bias in T cell responses due to a different representation of HLA class I molecules among the different cohorts, we characterized the HLA class I haplotype of each participant via either genomic sequencing or HLA-specific antibody staining, according to sample availability. For the HLA class I molecules included in the SARS-CoV-2 epitope prediction, the coverage among the four cohorts was at comparable levels for the majority of HLA class I molecules (Supplementary Fig. 4).

Except for pre-pandemic donors, blood was collected over several months, thus allowing us to investigate the longevity of SARS-CoV-2 CD8^+^ T cell immunity. We first looked at CD8^+^ T cell responses and antibody titers at early time points after diagnosis (PCR test) for mild COVID-19 and after enrollment in the study for asymptomatic seropositive cohorts. We observed concomitantly detectable levels of SARS-CoV-2-specific IgG and CD8^+^ T cells in the majority of the individuals (Fig. 1D), in line with previous reports describing coordinated responses of the humoral and T cell arms in individuals who resolved the infection without the development of severe symptoms^4^. Despite the fact that antibody titers rapidly waning over time (Fig. 1E), SARS-CoV-2-specific CD8^+^ T cells remained relatively stable (Fig. 1F), similar to other reports^8,20^, and, remarkably, were detected up to twelve months post-infection (Fig. 1G). Overall, we could show that the designed 9-mer pool can be used to detect SARS-CoV-2 CD8^+^ T cell responses regardless symptom severity, and that long-lasting CD8^+^ T cell immunity establishes upon primary infection.

### Identification of immunodominant SARS-CoV-2 CD8^+^ T cell epitopes

Before investigating the functionality of SARS-CoV-2-specifc TCR repertoires, we searched in our 9-mer pool for immunodominant epitopes, in order to study CD8^+^ T cell responses specific for single epitopes that might have high relevance in SARS-CoV-2 infection.

To do so, we applied a two-step deconvolution process. First, the 9-mer pool was split into four distinct sub-pools, each composed of 8-12 peptides, which we used to re-stimulate PBMCs previously expanded on the entire 9-mer pool (Supplementary Fig. 5). In a second step, peptides from reactive sub-pools were tested individually on the same donors. We performed this deconvolution on mild COVID-19 responders (Fig. 1C) for whom HLA class I genotype was available (n = 34). We found 19 immunogenic peptides eliciting IFN-γ secretion in CD8^+^ T cells, with a certain variability in the magnitude of responses and the in number of responders (Fig. 2A and Supplementary Fig. 6). Except for ORF1_LTN and ORF1_HSI, immunogenicity of the remaining epitopes was reported also in other studies^8,24,27,33,35,36^ (Supplementary Table 1). Interestingly, we often observed CD8^+^ T cell responses against multiple epitopes deriving from the same SARS-CoV-2 ORF (e.g. donor #11 and #12) as well as from different ORFs (e.g. donor #14 and #24) (Fig. 2A) in individual donors, indicating that broad and polyclonal CD8^+^ T cell response are elicited upon infection.

**Figure 2.**
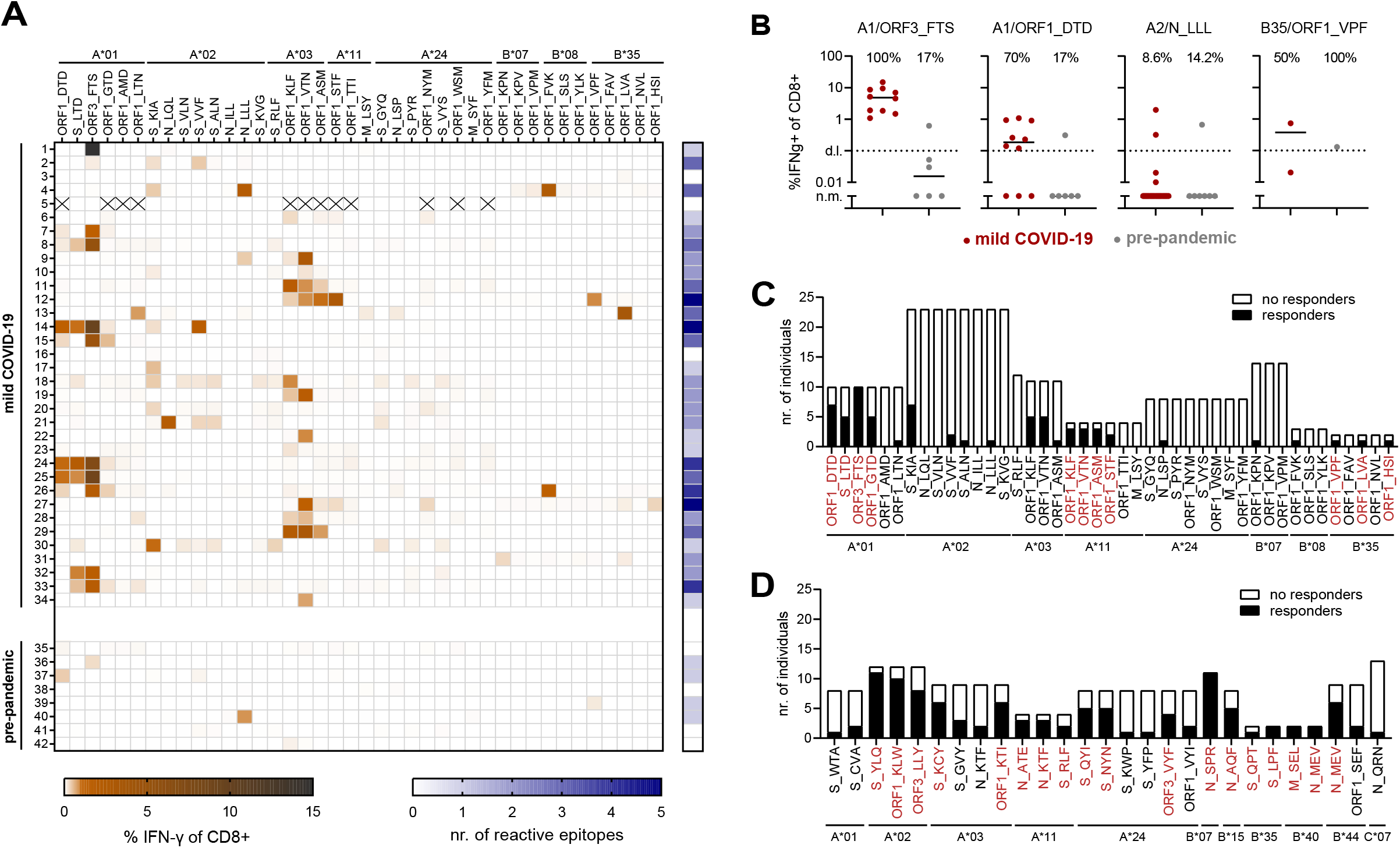
Identification of immunodominant SARS-CoV-2-specific CD8^+^ T cell epitopes. PBMCs from mild COVID-19 (n = 34) or pre-pandemic donors (n = 8) were *in vitro* expanded via co-culture with autologous PBMCs pulsed with 10 µg/µl 9-mer peptide pool for 10-12 days and re-restimulated with one of four peptide subpools (1 µg/µl). According to subpool responses, reactivity to individual peptides (1 µg/µl) was conclusively tested. **A)** Heat map showing the percentage of CD8^+^ T cells producing IFN-γ in response to individual peptide stimulation (orange-black gradient scale) and the number of reactive epitopes per donor (blue-white gradient scale) of mild COVID-19 (Donor 1-34) and pre-pandemic donors (Donor 35-42). **B)** Comparison of CD8^+^ T cell responses after *in vitro* expansion to specific epitopes in mild COVID-19 (red) and pre-pandemic (grey) individuals. Percentages indicate immunodominance. **C-D)** Immunodominance of individual SARS-CoV-2 epitopes in mild COVID-19 for peptides of the in-house 9-mer peptide pool (C) and subsequent peptide selection to increase HLA coverage (D). Peptides inducing > 50 % response rate in HLA-matched donors are marked in red. For flow-cytometry analyses, reactive cells were pre-gated on CD3^+^ living lymphocytes.

We also assessed the specificity of the identified immunogenic epitopes to SARS-CoV-2 through evaluation of single-peptide responses in the few pre-pandemic responders. We confirmed CD8^+^ T cell reactivity and identified immunogenic epitopes only for four of them. In addition, we observed responses to only one epitope per donor and at low magnitude (Fig. 2A, donors 35-42), overall highlighting how less consistent and robust these responses are comparable to mild COVID-19. All four epitopes stimulating CD8^+^ T cells in pre-pandemic individuals were found immunogenic also in mild COVID-19 (Fig. 2A). Despite a similar frequency in the unexposed individuals (except for B35/ORF1_VPF), ORF1_DTD and ORF3_FTS epitopes showed remarkably high immunodominance in mild COVID-19 (Fig. 2B), which could be explained by an unusually high precursor naïve frequency similar to the findings of Nguyen et al.^34^ In contrast, similar percentages of responders were found in mild COVID-19 and unexposed for A2/N_LLL and B35/OFR1_VPF epitopes (Fig. 2B), which would more support the hypothesis of cross-reactive epitopes with limited relevance in SARS-CoV-2 infections. Additional analyses would be necessary to decipher comprehensively the source of pre-pandemic responses.

Next, to understand the relevance of the identified immunogenic epitopes in SARS-CoV-2 T cell immunity, we quantified their immunodominance by contextualizing CD8^+^ T cell responses with regard to the HLA background of the donors (in other words, we analyzed the number of responders to HLA-matched predicted epitopes). Among the 19 immunogenic SARS-CoV-2 epitopes, eleven showed immunodominance of at least 50% (Fig. 2C, marked in red). By combining the responses of individual epitopes restricted to the same HLA class I molecule, we achieved a response rate of 100% for HLA-A*01:01, HLA-A*11:01 and B*35:02 and 63% for HLA-A*03:01 but only 40% for HLA-A*02:01, 33% for HLA-B*08:01 and less than 15% or no responses for the remaining HLAs (Supplementary Fig. 7A). To increase HLA coverage, we further tested a second pool of SARS-CoV-2-derived 9-mers composed of a mixture of newly predicted and published epitopes (Supplementary Table 1). We confirmed additional 27 immunogenic epitopes, of which 17 displayed an immunodominance higher than 50% (Fig. 2D and Supplementary Fig. 7B); furthermore, we sharply increased the response rate for HLA-A*02:01 (70%), A*24:02 (78%), B*07:02 (100%) and gained coverage for HLA-B*40:01 (100%) and B*44:03 (60%) (Supplementary Fig. 7B.

Overall, we identified a pool of immunodominant SARS-CoV-2 epitopes that could specifically identify individuals who had been exposed to the virus. Indeed, despite sporadic responses in non-infected individuals, the unique pattern of multiple-epitope responses makes convalescent COVID-19 individuals uniquely distinguishable.

### Isolation of SARS-CoV-2-specific MHC class I-restricted TCRs

Previous analyses have revealed several promising CD8^+^ T cell epitopes with high immunodominance and specificity to SARS-CoV-2, but without providing any information on the quality of the detected T cell responses. To fill this gap in knowledge, we identified and functionally characterized SARS-CoV-2 epitope-specific TCR repertoires.

We first focused on two highly immunodominant SARS-CoV-2 epitopes restricted to frequent HLA class I molecules (A1/ORF1_VTN and A3/ORF3a_FTS). PBMCs from HLA-matched, mild COVID-19 individuals were expanded on the 9-mer pool and re-challenged with either A1/ORF1_VTN or A3/ORF3a_FTS epitope prior to flow-cytometric cell sorting followed by single-cell RNA sequencing (scRNA-seq). In addition to CD8^+^IFN-γ^+^ T cells, which should be enriched in freshly re-activated T cells specific for the investigated epitopes, we also sorted CD8^+^IFN-γ ^-^ T cells where TCRs specific for either other immunogenic epitopes of the 9-mer pool or completely unrelated pathogens could be found (Fig. 3A). In this way, on the one hand, most of the information about SARS-CoV-2-specific TCR repertoire of a donor would be retrieved without losing the focus on the epitopes of highest interest. On the other hand, the potential of gene signatures to infer TCR specificity and functionality may be explored. In total, four donors (two donors for each selected peptide) were investigated (Supplementary Fig. 8). Analysis of clonotypic expansion and Leiden clustering revealed that the most expanded TCR clonotypes were present in clusters 0, 1 and 5 (Fig. 3B-C), indicating that these three clusters may contain activated and expanded SARS-CoV-2-specific T cells. Particularly cluster 1 contained cells with high, and for some markers unique, expression of effector molecules (*IFNG, GZMB* and *IL-2*) and activation markers (*XCL1, CD69 and CRTAM*) (Fig. 3D). The XCL1 chemokine is produced by activated T cells during infections and inflammatory responses and interacts with XCR1 receptor on dendritic cell, thereby promoting dendritic cell-mediated cytotoxic immune response^37^. CRTAM is expressed on activated T cells^38,39^ and coordinates cell polarity during activation, which was shown crucial for production of effector cytokines^40^. Together with the up-regulation of *IFNG, GZMB, IL-2* and *CD69* genes, this signature of recent activation may suggest an enrichment of T cells specific for A1/ORF1_VTN and A3/ORF3a_FTS epitopes in cluster 1.

**Figure 3.**
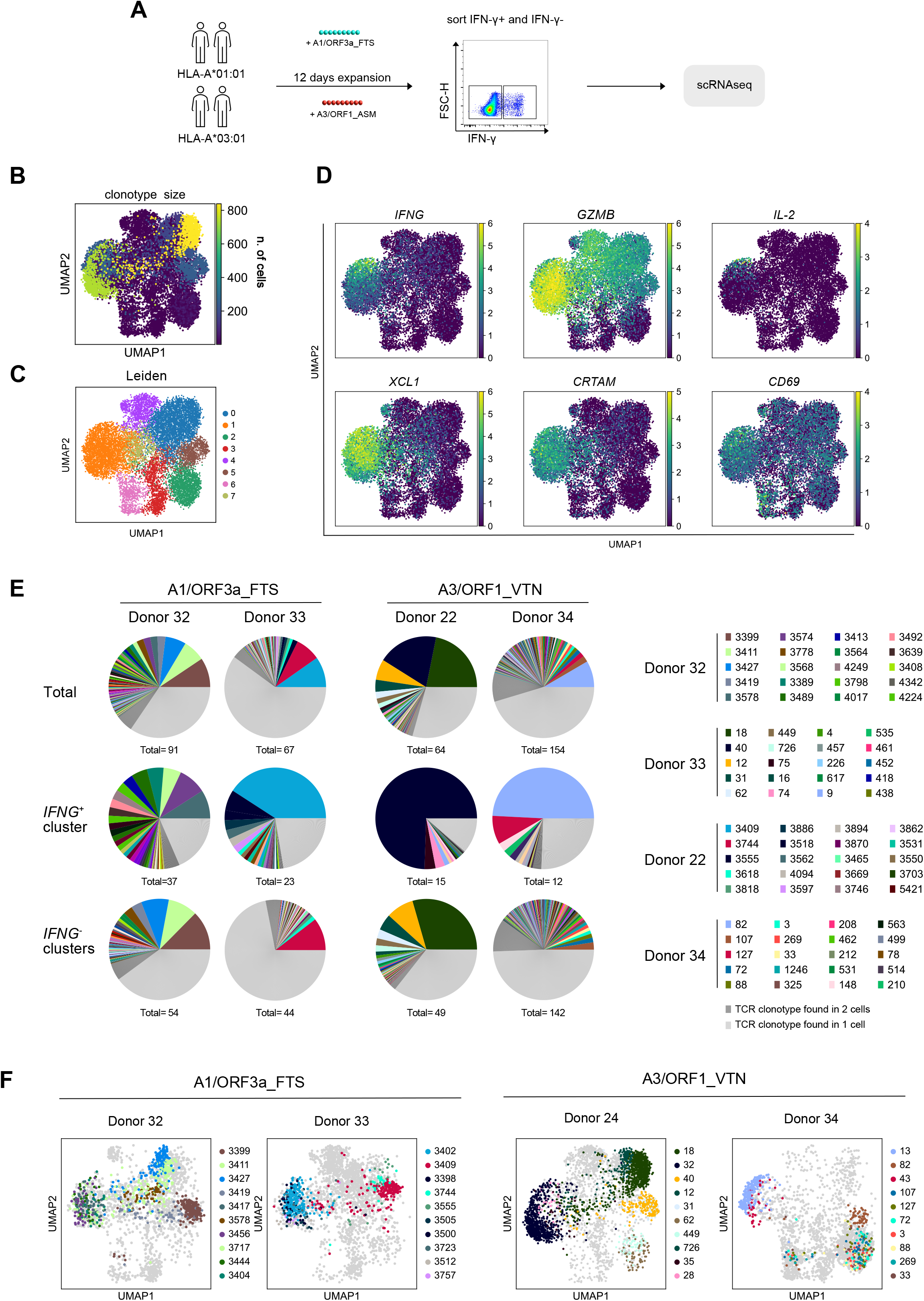
Isolation of SARS-CoV-2-specific TCRs. **A)** Schematic depiction of the strategy for isolating SARS-CoV-2-specific TCRs. Briefly two HLA-A*01:01 and two HLA-A*03:01 mild COVID-19 donors were *in vitro* expanded via co-culture with autologous PBMCs pulsed with 10 µg/µl 9-mer peptide pool for 10-12 days, and freshly re-stimulated with 1 µg/µl crude A1/ORF1_VTN and A3/ORF3a_FTS epitopes, respectively, for 4 h prior to sorting. For each donor, 2.500 CD8^+^IFN-γ^+^ and 10.000 CD8^+^IFN-γ^-^ T cells were sorted. Donors with same HLA background were pooled prior processing for single-cell RNA sequencing. **B)** UMAP neighborhood embedding showing distribution of TCR clonotypes and corresponding sizes. **C)** Depiction of Leiden clustering (resolution=0.5) according to the neighborhood embedding. **D)** UMAP neighborhood embedding showing distribution of T cell function and activation markers. **E)** Pie chart showing the percentage of each TCR clonotype of the total repertoire (top), Leiden cluster 1 (middle) and all Leiden clusters except of 1 (bottom). **F)** Pooled samples were demultiplexed by single nucleotide polymorphisms (SNPs) and assigned to a specific donor according to HLA class I haplotype. For each donor, distribution of the 10-top TCR clonotypes within the neighborhood embedding is depicted.

Pooled samples were deconvoluted according to single nucleotide polymorphisms^41^ and assigned to an individual donor by gender and HLA class I genotype (Supplementary Fig. 9 A-C). All donors showed a polyclonal TCR repertoire (Fig. 3E) with the dominating clonotypes distributed among clusters 0, 1 and 5 (Fig. 3F). The TCR repertoire for cluster 1 (*IFNG*^+^ cluster) was highly diverse, in particular in case of ORF3a_FTS and showed a higher fraction of clonally expanded TCRs (Fig. 3E).

In summary, we found that SARS-CoV-2-specific TCR repertoires are highly polyclonal with some clonotypes showing a prominent signature of recent activation, presumably reflecting fresh re-stimulation.

### SARS-CoV-2-specific TCR repertoires contain highly functional TCRs

To investigate the quality of the identified SARS-CoV-2-specific TCR repertoires, we next re-expressed candidate TCRs for functional characterization. We selected TCRs from both cluster 1 (TCR 13, TCR 28, TCR 32 and TCR 43 from HLA-A*03:01 donors and TCR 3398 and TCR 3456 from HLA-A*01:01 donors) and clusters 0 and 5 (TCR 18 and TCR 82 from HLA-A*03:01 and TCR 3399 and TCR 3409 from HLA-A*01:01 donors) among the top 10 expanded clonotypes (Fig. 3F and Supplementary Fig. 9D). Thus, potential associations between TCR specificity/functionality and the observed activation signature could be analyzed. Transgenic TCRs were introduced into primary T cells by CRISPR/Cas9-mediated orthotopic TCR replacement (OTR), where complete replacement of the endogenous TCR is achieved by the knock-in of the transgenic TCR into the endogenous TCR α locus and the concomitant knock-out of the TCR β locus^42^. Eventually, we generated TCR-transgenic T cells with near-physiological features that recapitulate the original antigen-specific T cell as closest as possible.

We first tested the specificity of our transgenic TCRs against ORF1_VTN and ORF3a_FTS epitopes via epitope-HLA multimer staining. All TCRs selected from cluster 1 (*IFNG*^+^ cluster) showed strong staining toward the corresponding HLA-matched multimer (relevant multimer), except for A3/TCR 32 that stained weaker. No staining was instead observed for irrelevant multimers i.e. HLA-A*03:01 or HLA-A*01:01 multimer loaded with a different epitope (SARS-CoV-2 ORF1_KLF and pp50_245-253_, respectively), thus confirming specific epitope recognition (Fig. 4A-B). Instead, none of the TCRs selected from clusters 0 and 5 (*IFNG*^-^ cluster) reacted to ORF1_VTN and ORF3a_FTS multimers, but A3/TCR 40 showed specificity for ORF1_KLF (Fig. 4A-B). To evaluate whether the remaining *IFNG*^-^ clusters-derived TCRs could recognize SARS-CoV-2 epitopes at all, we pulsed autologous PBMCs (from the same donor the TCR was isolated from) with SARS-CoV-2 9-mer pool and used them to stimulate TCR-engineered T cells; cytokine release was observed only for A1/TCR 3399 (Supplementary Fig. 10). As CMV and EBV infections have high prevalence in the human population and usually induce big-size clonal responses^43,44^, we also stimulated our engineered T cells with CMV and EBV peptide pools but no cytokine release was observed (Supplementary Fig. 10). Altogether, HLA multimer staining confirmed that cluster 1 was enriched in freshly re-stimulated TCRs and that signature of recent activations are indicative of TCR specificity to recent epitope re-challenge. TCRs with clear specificity for SARS-CoV-2 epitopes were further characterized in terms of functionality by means of cytokine release and cytotoxicity upon peptide stimulation. All TCR-engineered T cells responded to the cognate peptide stimulus in a dose-dependent manner, and the majority showed sensitivity to very low peptide concentrations, as indicated by the IFN-γ EC_50_ values. A3/TCR 13, A3/TCR 28, A1/TCR 3398 and A1/TCR 3456 showed particularly high functionality with half-maximum cytokine release at peptide stimulation above 10^−7^ M, whereas A3/TCR 43 and A3/TCR 32 showed intermediate and low functionality, respectively (Fig. 4C-D).

**Figure 4.**
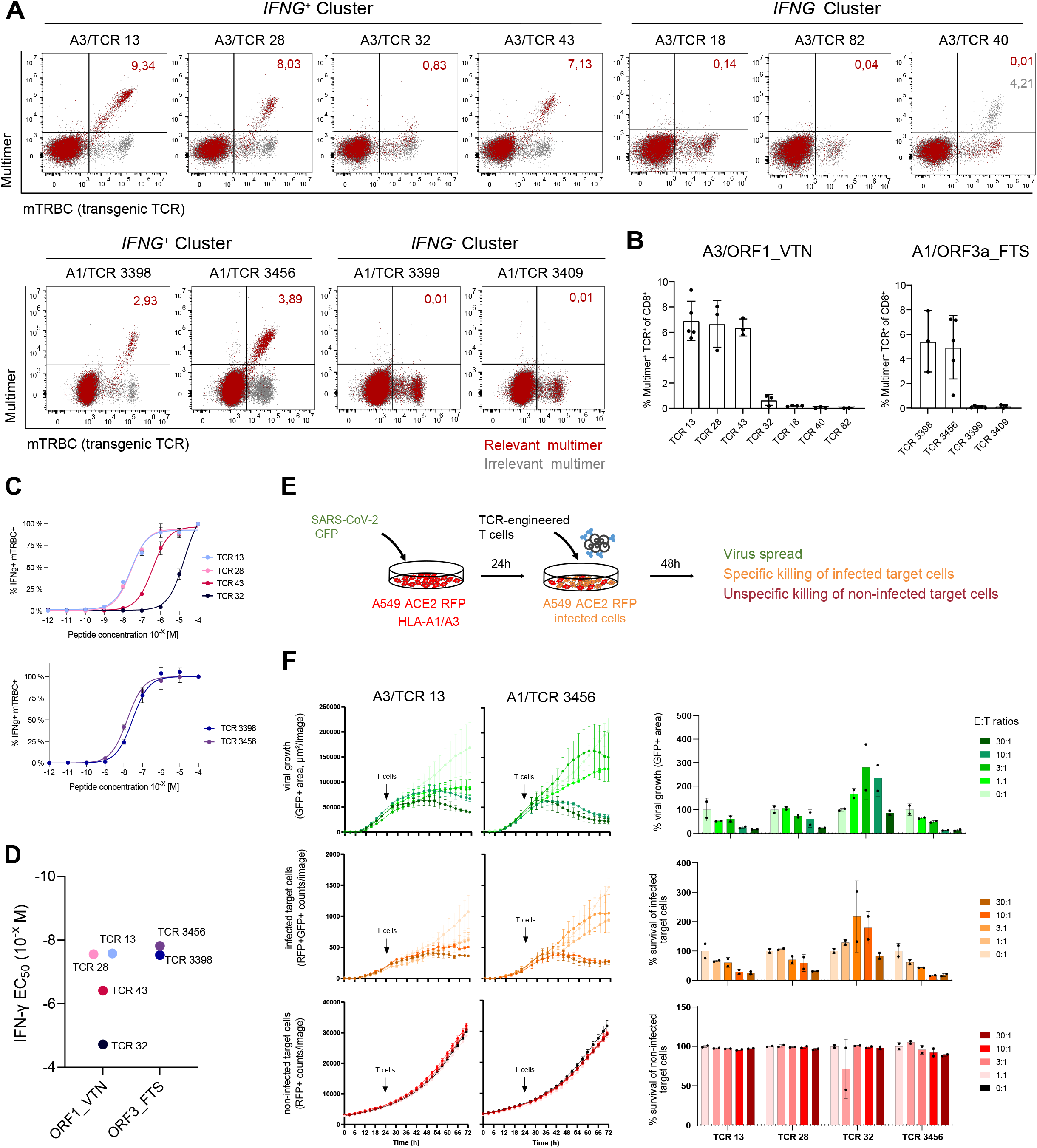
Identification of highly functional and cytotoxic SARS-CoV-2-specific TCRs. PBMCs from healthy donors were engineered to express a transgenic TCR, which contained a murine constant region, via CRISPR/Cas9-mediated orthotopic replacement. **A-B)** TCR-engineered T cells were stained with pMHC multimers loaded with either A1/ORF1_VTN or A3/ORF3a_FTS epitopes (relevant multimer, shown in red), in addition to anti-CD8 and anti-mTRBC (constant region of the murine β chain) antibodies. As control, multimers loaded with A1/pp50_245-253_ or A3/OFR1_KLF (irrelevant multimers, shown in grey) were used. Representative examples of multimer staining with indicated frequencies of multimer^+^ TCR-KI^+^ of CD8^+^ cells (A). Quantification of multimer^+^ TCR-KI^+^ of CD8^+^ cells (B). Cells were pre-gated on CD8^+^ living lymphocytes. **C)** Intracellular IFN-γ staining of T cells engineered with SARS-CoV-2-specific TCRs after 4 h stimulation with K562 antigen-presenting cells expressing either HLA-A*01:01 or HLA-A*03:01 molecules and pulsed with the indicated peptide concentrations. **D)** IFN-γ EC_50_ of SARS-CoV-2 TCR-engineered T cells. **E)** A549 cells were retrovirally transduced to express ACE2, a red fluorescent protein (RFP) and an HLA class I molecule of interest. Target cells were infected with a GFP-expressing SARS-CoV-2 virus and, 24 h post infection, co-cultured with sorted TCR-engineered CD8^+^ T cells at different effector-to-target ratios (E:T) for additional 48 h. As control, engineered T cells were added also to non-infected cells. **F-G)** Live tracking (F) and end-point quantification (G) of viral spread (GFP^+^), infected cells (GFP^+^RFP^+^) and non-infected cells (RFP^+^). For end-point analyses, infected cells were washed before signal acquisition.

To test for cytotoxicity, we generated a target cell line susceptible to SARS-CoV-2 infection. Briefly, the A549 human lung cancer cell line was engineered to express ACE2 protein, red fluorescent protein and HLA-A*01:01 or HLA-A*03:01 molecules, which allowed viral infection, live-cell imaging acquisition and appropriate epitope presentation, respectively. A genetically modified GFP-expressing SARS-CoV-2 virus was used for infection. 24h after viral infection, addition of TCR-engineered T cells successfully blocked viral spread (GFP+ signal) and induced killing of infected target cells (RPF+GFP+) with minimal off-target toxicity on non-infected cells (RFP+) (Fig. 4E-F). Only A3/TCR 32 showed again extremely low functionality, in line with the low levels of cytokine release upon epitope re-stimulation.

In summary, we showed that the TCR repertoire specific for SARS-CoV-2 ORF1_VTN and ORF3a_FTS epitopes contains highly functional and cytotoxic TCRs and that expression of genes related to recent activation is indicative of epitope specificity.

### Functional SARS-CoV-2-specific TCRs are recruited in non-severe SARS-CoV-2 infection

The validated TCRs compose a minor part of the total repertoire we isolated out of ORF1_VTN and ORF3a_FTS specific T cells. Expression of *IFNG*, among other genes, was indicative of epitope specificity, as it reflected reactivity after fresh re-stimulation prior to sorting and sequencing. Therefore, we searched our transcriptomic dataset for gene signatures that could associate with T cell functionality in order to accurately predict functionality also for non-validated TCRs.

Taking into consideration that TCRs were isolated from *in vitro* expanded CD8^+^ T cells after fresh re-stimulation, we analyzed the predictive potential of an available gene signature related to CD8^+^ T cell activation. CD8^+^ activation score was enriched in cells expressing freshly re-stimulated TCRs, but it showed only a trend of correlation with T cell functionality among the reactive TCRs (IFN-γ EC_50_) (Fig. 5A). To improve the sensitivity of prediction, we defined two additional gene signatures more specific for our dataset and based on gene expression of cells expressing the TCRs that we selected for re-expression and characterization; a “reactivity signature” composed of genes differentially expressed between epitope-reactive TCRs and non-reactive TCRs (Supplementary Fig. 11A), and a “functionality signature” comprising genes that best correlated with IFN-γ EC_50_ (Supplementary Fig. 11B-C). Epitope-reactive TCRs showed high scores for both signatures and, remarkably, functionality score accurately predicted T cell functionality. The percentage of *IFNG*^+^ cells, but not the clonotype size, also clearly identified epitope-reactive TCRs but was not sufficient to resolve low (TCR 32) and highly functional TCRs (TCR 13 and TCR 28 among others) (Fig. 5A). When applied to the entire identified TCR repertoires, high activation and functionality scores clearly separated and identified SARS-CoV-2-specific T cells responding to the recent re-stimulation, which were almost all belonging to the *IFNG*^+^ cluster (Fig. 5B). More importantly, within the reactive TCRs, clonotypes distributed among a range of signature scores. We found a relevant proportion of TCRs with scores similar to the high-avidity TCR 3456, TCR 3398, TCR 28 and TCR 13, except for Donor 22 for whom higher amount of low functional TCRs was observed. Intriguingly, low-avidity TCR 32 correspondingly showed an intermediate positioning in the activation/proliferation score landscape (Fig. 5B). Overall, our data indicate that a polyclonal population of highly functional TCRs is recruited in mild COVID-19, despite a variable proportion of intermediate and low avidity TCRs.

**Figure 5.**
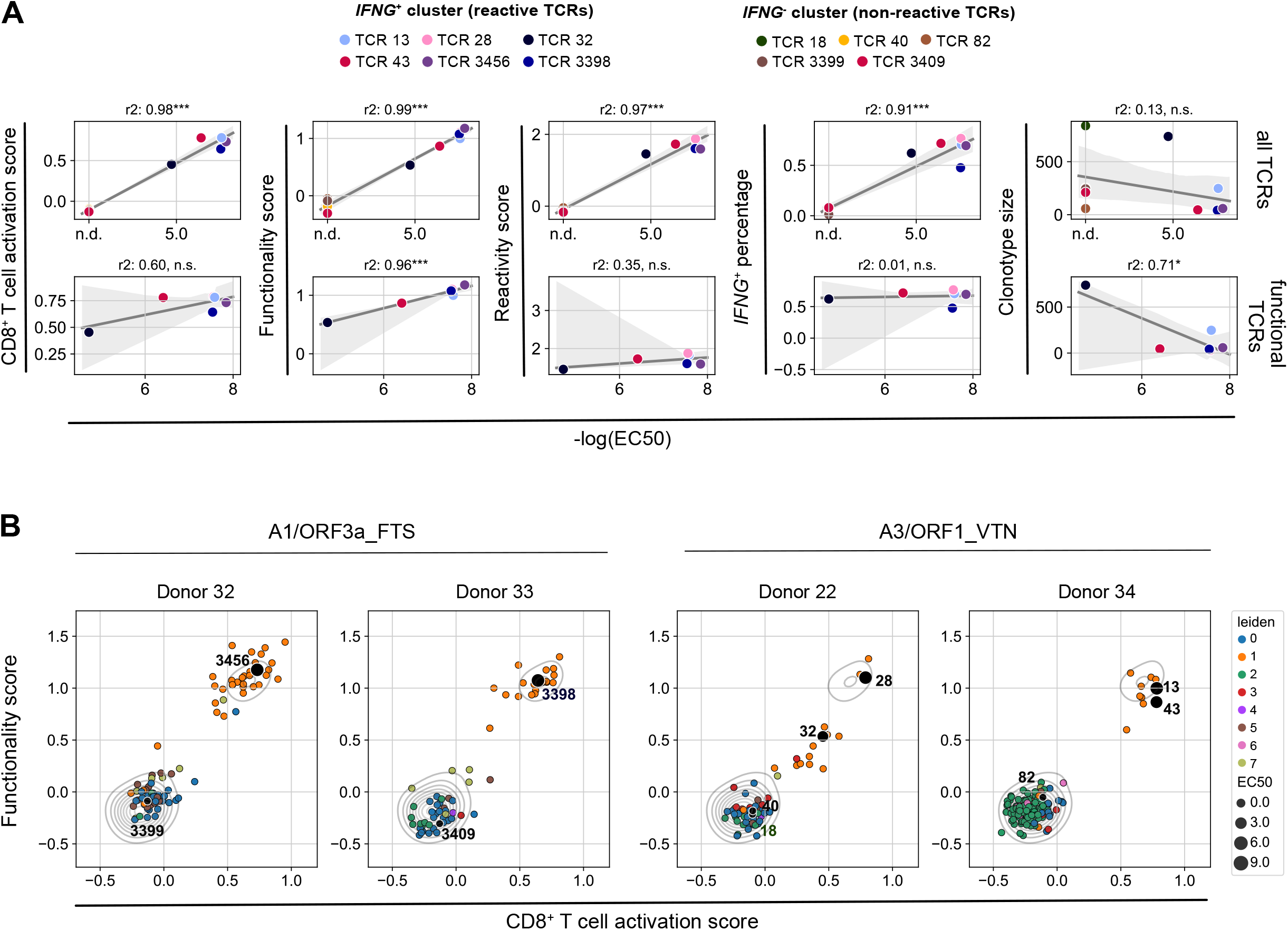
Highly functional TCRs correlate with signature of recent activation. **A)** Linear regression analysis between *in vitro* functionality (IFN-γ EC_50_) and gene scores for all (upper row) and functional (lower row) TCRs. The gray area depicts the 95% confidence interval. * p value < 0.05, ** p value < 0.01, *** p value < 0.001, **** p value < 0.0001. Two-sided p-values for a hypothesis test (null hypothesis: slope is zero) have been calculated in (j-l), using Wald Test with t-distribution of the test statistic. **B)** TCR clonotype distribution according to CD8^+^ T cell activation score and functionality score. The color code refers to the Leiden clusters and the dot size of the re-expressed TCRs corresponds to the IFN-γ EC_50_ value. Contours show Kernel-density estimates (KDE) of the pooled dataset including the four donors depicted in the individual plots.

We finally expanded this TCR repertoire analyses to additional nine immunodominant SARS-CoV-2 epitopes (Fig. 2D) restricted to five different HLA class I molecules in eight mild COVID-19 individuals. Each donor was stimulated with a specific epitope of interest and expanded *in vitro* prior to re-stimulation and sorting of CD8^+^IFN-γ^+^ cells. Each individual was additionally labeled with a DNA-tagged antibody in order to make it distinguishable when pooled with other donors for scRNA-seq (Fig. 6A). Despite sorting, transcriptomic data revealed heterogeneous expression of genes related to T cell function and activation, which resulted in different enrichments in the reactivity, functionality and activation scores (Fig. 6B). High reactivity scores were broadly observed, suggesting that the majority of the isolated repertoire should be specific to SARS-CoV-2. In addition, a fraction of those TCRs was particularly enriched also in the functionality score, as well as in the activation score, indicating the presence of TCRs of presumably high functionality (Fig. 6B). For a better evaluation, we correlated the reactivity and functionality scores for each clonotype of the newly identified repertoires, and we used the *in vitro* characterized TCRs (Fig. 4-5) as controls. We selected the functionality score over the activation score due to its higher accuracy in predicting TCR functionality (Fig. 5A). We observed a bimodal distribution, with the functional transgenic TCRs (TCR 13, TCR 28, TCR 43, TCR 3456 and TCR 3398) occupying the score^high^ cluster and the low-avidity (TCR 32)/no-specific TCRs (TCR 18, TCR 40, TCR 82, TCR 3409 and TCR 3399) showing low scores. All other clonotypes distributed within the reactivity/functionality landscape and a relevant number of clonotypes overlaid with highly functional transgenic TCRs (Fig. 6C), thus further corroborating our initial observations.

**Figure 6.**
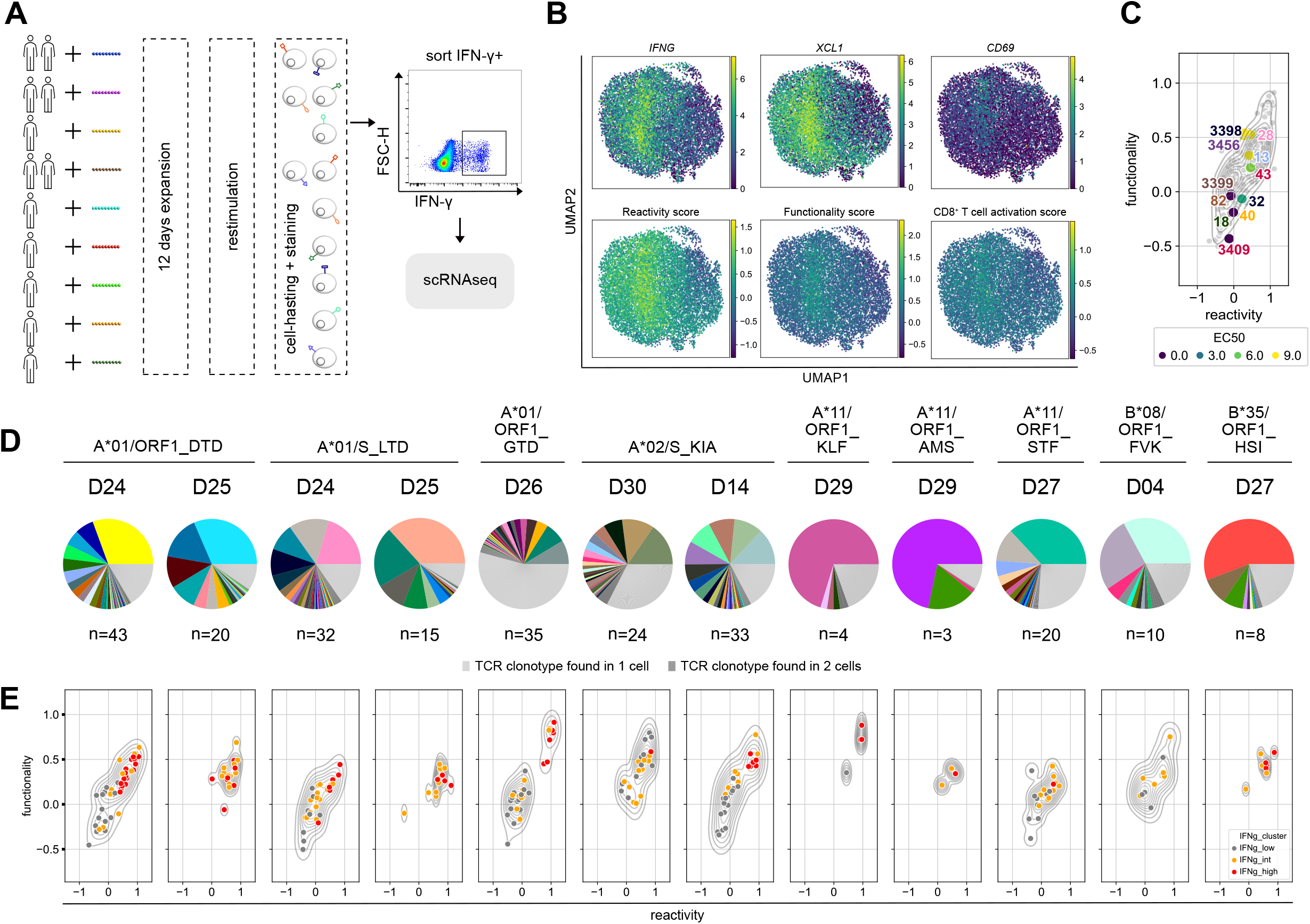
Functional TCRs are recruited in non-severe SARS-CoV-2 infections. **A)** Schematic depiction of the experimental setup. **B)** UMAP neighborhood embedding showing distribution of T cell function and activation marker expression across the dataset. **C)** TCR clonotype distribution according to reactivity score and functionality score. Functionally validated TCRs from Figure 5 were overlaid as benchmark. Contours show Kernel-density estimates of the dataset whereas dot color corresponds to the IFN-γ EC_50_ of the validated TCRs. **D)** Pie chart showing the percentage of each TCR clonotype of the total repertoire for the individual peptides and donors. **E)** TCR clonotype distribution according to reactivity score and functionality score for the individual peptides and donors. Contours show Kernel-density estimates (KDE) for each individual sample, the color code corresponds to the IFNG expression batches (low, intermediate, high).

As a next step, we wanted to understand the functional landscape of epitope-specific TCR repertoire. To do so, we deconvoluted the two sample pools and assigned each sample to one donor according to DNA barcodes (Supplementary Fig. 12). Most of the donors showed again a highly polyclonal TCR repertoire (Fig. 6D). Remarkably, highly functional (defined by high gene signature scores) TCRs were predicted for all donors and analyzed SARS-CoV-2 epitopes (Fig. 6E).

Overall, we could show that polyclonal CD8^+^ T cell responses are elicited against immunodominant SARS-CoV-2 epitopes and that highly functional TCRs are recruited in mild COVID-19, despite some variability according to HLA-epitope combination.

## DISCUSSION

We investigated the functional landscape of MHC class I-restricted TCR repertoires specific for SARS-CoV-2 in the context of non-severe infections, where protective immunity should develop. SARS-CoV-2-specific TCRs were isolated based on cytokine release after short *in vitro* peptide stimulation. T cells genetically engineered with those TCRs showed heterogeneous functionality, where highly functional TCRs efficiently killed virus-infected target cells and inhibited viral spread. T cells expressing functional TCRs were enriched in gene signatures of recent activation, which were retrieved for each epitope:HLA combination analyzed, thus indicating that highly functional TCRs are commonly recruited in non-severe SARS-CoV-2 infections.

The importance of CD8^+^ T cells in respiratory virus infections is well recognized. CD8^+^ T cells are recruited to the lungs within 8-10 days from infection^45–47^, contribute to viral clearance^48–50^, and generate a pool of memory cells that protect from re-infection^46,48^. Also in SARS-CoV infections, CD8^+^ T cells provided substantial protection in preclinical studies^51^ and long-lasting memory SARS-CoV-specific CD8^+^ T cells have been detected up to 17 years after infection in humans^24^. T cell immunity against SARS-CoV-2 shares many of the above-mentioned aspects. Early recruitment of T cells prevented severe disease^15^ and was usually followed by the establishment of a robust pool of functional memory T cells^6,27^ detectable up to 6-10 months after infection^20,52–54^. In addition to this existing body of evidence, we further showed that SARS-CoV-2-specific CD8^+^ T cells are detectable up to 12 months from infections, a hint pointing towards long-lasting immunity similar to SARS-CoV. Of course, continuous follow-up is necessary to strengthen the interpretation of even longer maintenance of CD8^+^ T cell immunity.

However, whether and how CD8^+^ T cells may mediate protective immunity in SARS-CoV-2 infections needs to be investigated in more detail. Despite encouraging preclinical data showing loss of protection following CD8^+^ T cell depletion^55^, evidence of the protective role of CD8^+^ T cells in humans is still scarce. SARS-CoV-2-reactive CD8^+^ T cells have often been functionally characterized using *ex vivo* antigen stimulation-based assays (i.e. IFN-γ ELISpot assays, intracellular cytokine staining, and activation-induced marker), but the use of a single peptide (or peptide mix) dose hinders discrimination of high and low functional T cells and thus obscures the actual range of functionality. T cell functionality is primarily encoded in its TCR. For this reason, as a first step towards a deeper understanding of the quality of CD8^+^ T cell responses against this new virus, we investigated the functionality of SARS-CoV-2-specific TCR repertoires in individuals who recovered from mild symptomatic disease. First, we learned that in this clinical situation, SARS-CoV-2 epitope-specific CD8^+^ T cells are very low in frequency in peripheral blood, making it difficult to establish a direct *ex vivo* characterization assay from a limited sample size. However, a short step of peptide-mediated *in vitro* expansion^32^ allowed us to robustly detect SARS-CoV-2-specific CD8^+^ T cells in convalescent individuals, similar to other reports^27,33,56^.

By combining this *in vitro* peptide expansion with single-cell RNA sequencing, we accessed the MHC class I-restricted TCR repertoire specific towards immunodominant, and thereby relevant for the course of the infection, SARS-CoV-2 epitopes. Beside validation of SARS-CoV-2 specificity and peptide sensitivity of selected transgenic TCRs after re-expression in primary T cells, of particular importance and uniquely from other studies^57^ was the experimental validation that TCRs with high sensitivity towards SARS-CoV-2-specific epitopes can mediate cytotoxicity against virus-infected cells. The association between immunogenicity (the ability of eliciting an immune response) and protection (the direct contribution to the resolution of infection by the elicited immune response) is not self-explanatory. Immunogenic epitopes can be efficiently cross-presented but eventually not expressed in infected target cells^58,59^, in particular for *in silico* predicted epitopes^60^, thereby inducing T cell responses that are neither relevant nor indicative of the course of the infection. For two epitopes (A1/ORF1_VTN or A3/ORF3a_FTS), we showed for the first time that T cells genetically engineered with SARS-CoV-2-specific TCRs were capable of directly killing virus-infected cells *in vitro* through specific epitope recognition, as non-infected cells were not recognized. This evidence gives an additional value to the epitopes under investigation. Besides immunogenicity and immunodominance, which has been widely corroborated by other studies where the landscape of HLA class I-restricted SARS-CoV-2 epitopes was exhaustively deciphered^21^, the epitopes we described in the study are indicative of recruitment of highly functional CD8^+^ T cell responses. Thus, they represent a particularly useful tools for studying the quality of T cell responses in several contexts e.g. after vaccination and severe infections. The identification of functional and highly effective SARS-CoV-2-specific TCRs also opens up possibilities for therapeutic use based on TCR-engineered T cells in individuals with high risk of developing a severe clinical course. The therapeutic value of adoptive transfer of antigen-specific, TCR-engineered T cells is well recognized^61,62^, especially now that precise genetic engineering offers the flexibility of generating increasingly sophisticated but near-to-physiological autologous T cell products^42^. Furthermore, accumulated evidence supports a model where late recruitment of T cells^15,63^, presumably due to a delayed activation of type I interferon response^64^, contributes to disease progression, thus offering a therapeutic window for the adoptive transfer of potentially curative TCR-engineered T cells. Recent findings also disproved initial concerns about the potential role of T cells in immunopathology in advanced COVID-19 diseases, rather attributed to the massive amount of lung-infiltrating neutrophils and circulating monocytes^65^.

A limitation of our study is the use of an *in vitro* expansion step prior T cell analyses, which may bias the relative abundance of clonotypes and may favor outgrowing of some clones at the expense of others. However, this approach of peptide stimulation offered the possibility of exploiting signatures of recent activation to discriminate epitope-specific and functional TCRs^66^. Indeed, we identified signatures of T cell function and recent activation correlating with TCR functionality, which allowed a broad *in silico* investigation of the functional landscape of entire TCR repertoires, otherwise unfeasible by conventional experimental validation. Remarkably, we predicted highly functional TCRs for each of the eleven immunodominant SARS-CoV-2 epitopes analyzed, despite a certain degree of functional heterogeneity. Together with the observed polyclonality, our data indicate that functional and diverse CD8^+^ T cell immunity should normally establish in non-severe SARS-CoV-2 infections. Polyclonality and high functionality are hallmarks of a protective TCR repertoire^67^, thus strongly supporting a similar role also in the context of SARS-CoV-2 infections. Nevertheless, the extension of TCR repertoire analyses to settings of severe infections remains a fundamental next step to assign CD8^+^ T cell responses a role as correlates of protection.

Overall, our data provide first evidence that SARS-CoV-2-specific CD8^+^ T cells persists up to 12 months after infection and are composed of a polyclonal and highly functional TCR repertoire capable of mediating direct killing of virus-infected cells. In addition, we provide tools – epitopes and TCRs – indicative of functional responses, useful for appropriately investigating and potentially diagnosing correlates of protection in severe patients.

## Data Availability

The manuscript contains all relevant data; beyond the main figures supplemantary data have been added to the manuscript..

## ACKNOWLEDGEMENTS

We thank Max Koch for the processing of blood for PBMCs isolation. We gratefully acknowledge Catharina Gerhards, Margot Thiaucourt and Laura Mirbach for their contribution in the logistic organization of blood sample delivery.

This study was supported by the EIT Health CoViproteHCt #20877, the German National Network of University Medicine of the Federal Ministry of Education and Research (BMBF; NaFoUniMedCovid19, 01KX2021; COVIM) and the DFG SFB1321 (Modeling and Targeting Pancreatic Cancer). E.D. was funded by the Corona-Forschungsanträge (Fakultät f. Medizin). V.G. and A. Pichlmair. were funded by ERC consolidator grant (ERC-CoG ProDAP, 817798), the German Federal Ministry of Education and Research (COVINET), and the German Research Foundation (PI 1084/5 and TRR179, TRR237) to AP. L.M.M. was supported by a PhD fellowship from the Boehringer Ingelheim Fonds.

## AUTHOR CONTRIBUTIONS

D.H.B., E.D. conceptualized the study; K.I.W. and L.M.M. performed experiments; K.I.W., L.M.M., S.J. and E.D. performed software analyses; S.J. and E.D. analyzed sequencing data; V.G. and A. Pichlmair. performed killing assays; H.P., H.B., S.S., S.H., A.S., V.H. and M.N. collected blood samples and clinical information of mild COVID-19; U.P., P.A.K., J.E., A. Priller, S.Y. and H.R. designed and organized the study on asymptomatic donors; M.O. and T.T. provided pre-pandemic donor PBMCs; K.W. performed HLA genotyping; T.B. and B.K. performed serology analyses; S.W., M.H. and H.M. contributed resources; D.H.B. and E.D. wrote the manuscript; K.I.W., L.M.M. and E.D. prepared figures; all authors read and approved the manuscript; D.H.B., M.G., C.S.C., K.S. and E.D. acquired the funding; D.H.B. and E.D. supervised the study and administered the project.

## RESOURCE AVAILABILITY

### Data and code availability

All data generated or analyzed during this study are included in this article, its supplementary information files and/or are available from the corresponding authors upon reasonable request. The notebooks containing all steps of data processing and analysis can be found online (https://github.com/SebastianJarosch/2021_Wagner_Mateyka_Jarosch_COVID_scRNAseq)

### Lead contacts

Elvira D’Ippolito or Dirk H. Busch, Institute for Medical Microbiology, Immunology and Hygiene, Technische Universität München (TUM), Munich, Germany

## EXPERIMENTAL MODELS AND METHOD DETAILS

### Clinical samples

For symptomatic SARS-CoV-2 infections, blood sample was collected at the Helios Klinikum München West, Deutsches Herzzentrum München and University Medicine Mannheim from healthcare employees who were diagnosed by PCR test and experienced mild symptoms (cold, cough and mild fever), for which home quarantine was sufficient. Participants donated 50 ml blood at the end of the required home quarantine and repeatedly in 4, 8, 12 and 24 weeks following recovery from infection. The study cohort of asymptomatic seropositive and seronegative donors was established at the Klinikum Recht der Isar (München) and included health care employees that were tested for presence of SARS-CoV-2 specific antibodies in April/May 2020. 10 ml blood was collected at two time points between August 2020 and November 2020. Frozen peripheral blood mononuclear cells (PBMCs) from pre-pandemic (unexposed) donors were received from the Institute for Transfusion Medicine Dresden and collected between 2018 and 2019. All participants provided informed written consent. Approval for the study design and sample collection was obtained within the framework of study “Establishment and validation of epitope-specific SARS-CoV-2 blood-based testing methods” (EPI-SARS) by the ethics committee of the Technical University of Munich (reference number 182/20) and the Institutional Ethics Committee of the University Medicine Mannheim (reference number 2020-556N).

### Epitope prediction

Potential CD8^+^ T cell epitopes were predicted from all open reading frames of the Wuhan-Hu-1 reference sequence (NC_045512) for MHC Class I binding to diverse HLA molecules (HLA-A*01:01, HLA-A*02:01, HLA-A*03:01, HLA-A*11:01, HLA-A*24:02, HLA-B*07:02, HLA-B*08:01, HLA-B*35:01) using NetMHC4.0 for peptides of 8 – 11 amino acids in length. Peptide candidates with predicted binding strength < 50 nM were further evaluated for immunogenicity, TAP transport, proteasomal cleavage and processing, using the following *in silico* prediction tools: Netstab1.0, NETCTL1.2, PickPocket1.1, NetMHCpan4.1. Peptide candidates showing the highest immunogenic prediction scores were cross-referenced for sequence homology to SARS-CoV-1, MERS and common cold corona viruses HCoV-OC43 (NC_006213), HCoV-HKU1 (NC_006577), HCoV-NL63 (NC_005831) and HCoV-229E (NC_002645). Peptides unique to SARS-CoV-2 with highest scores in various prediction tools were selected for further validation in an in-house peptide pool.

The peptide pool was supplemented with IEDB published SARS-CoV-2 epitopes and epitopes homologous to SARS-CoV-1 (NC_004718) and MERS (NC_019843) (Supplementary Table 1).

### Serology

For all time points of blood donation, a serum sample was taken and analyzed for anti-SARS-COV-2 IgG using the iFlash Immunoassay analyzer, following the manufacturer’s protocol. Briefly, the serum samples were incubated with samples treatment solution and SARS-CoV-2 antigen-coated paramagnetic microparticles to form a complex. Unbound material was washed from the solid phase and a second incubation step with Acridinium-labeled anti-human IgG conjugate followed. After washing, the pre-Trigger and Trigger solutions were added to the reaction mix and the resulting chemiluminescent reaction was measured as relative light units by the iFlash optical system. A cutoff was calculated from SARS-CoV-2 IgG calibrators.

### Cell culture

PBMCs were isolated from whole blood by gradient density centrifugation according to manufacturer’s instructions (Pancoll human) and frozen in fetal calf serum (FCS) + 10 % DMSO for liquid nitrogen storage. For T cell analyses, PBMCs were thawed and rested for 16 h in RPMI 1640 supplemented with 10 % FCS, 0.025 % l-glutamine, 0.1 % HEPES, 0.001 % gentamycin and 0.002 % streptomycin before stimulation or expansion procedures.

### T cell expansion with autologous peptide-pulsed PBMCs

20 % of total PBMCs were pulsed with 10 µg/ml peptide pool for 2 h at room temperature under gentle agitation at 1×10^6^ cells/ml. Excess of peptides was removed by washing and peptide-pulsed cells were co-cultured with the remaining 80 % of PBMCs for 10 - 12 days in RPMI 1640 supplemented with 5 % human serum, 0.025 % l-glutamine, 0.1 % HEPES, 0.001 % gentamycin and 0.002 % streptomycin at 1×10^6^ cells/ml. 50 IU/ml IL-2 was added every 3 - 4 days.

### T cell stimulation, cytokine release assay and T cell sorting

For intracellular cytokine release assay of *ex vivo* or post-expansion natural T cells, PBMCs were stimulated with 1 µg/ml peptide pool, 1 µg/ml peptide or 1 µg/ml PepTivator SARS-CoV-2 Protein S pool. For TCR-engineered T cells, K562 antigen presenting cells (retrovirally transduced with HLA-A1 or HLA-A3) were irradiated (80 Gy), loaded with different peptide concentrations (10^−12^, 10^−11^ 10^−10^, 10^−9^, 10^−8^, 10^−7^, 10^−6^, 10^−5^ and 10^−4^ M) overnight at 37 °C, and co-cultured with engineered T cells in 1:1 effector:target ratio. Incubation with peptides or antigen-presenting cells was done for 4 h at 37 °C, in presence of 1µg/ml GolgiPlug. DMSO served as negative control whereas 25 ng/ml PMA and 1 µg/ml Ionomycin served as positive control. After incubation, cells were stained with EMA solution (1:1000) for live/dead discrimination and subsequently with surface antibodies: CD19-ECD (1:100), CD8-PE (1:200), CD3-BV421 (1:100) and murine TCR β-chain-APC/Fire750 (1:100). Cells were fixed using Cytofix/Cytoperm solution followed by staining for intracellular cytokine by IFN-γ-FITC antibody (1:10). Flow cytometric analysis was performed on the CytoFlex S Cell Analyzer.

For cell sorting, expanded PBMCs (day 13 post-expansion) were freshly re-stimulated with 1 µg/ml peptide for 4 h at 37 °C. Peptide-reacting cells were sorted according to cytokine secretion, detected via IFN-γ catch (IFN-γ-FITC), and CD8 staining (CD8-eF450 1:200). Additionally, donor cells were stained with Total Seq-C antibodies (10x Hashtag 1-6, 0,5mg/ml) prior sample pooling, in order to discriminate individuals donors in the sequencing sample. Flow sorting of CD8^+^ IFN-γ^+^ and CD8^+^ IFN-γ^-^ was conducted on a MoFlo Astrios EQ under biosafety level 3.

### HLA genotyping

Generic PCR amplification of complete HLA-class I coding regions (HLA-A, -B and -C) were performed in a 11 µl polymerase chain reaction (PCR) assay using 5 µl LongAmp Taq 2x Master Mix, 3 µl forward and reverse primer mix (each 1.5 pmol/µl) and 3 µl DNA (about 10-50 ng/µl). The homemade amplification primers were located in the 5’- and 3’-untranslated region. On a SimpliAmp Thermocycler, the thermocycler profile used was a denaturation step at 94°C for 7 min, followed by 15 cycles of denaturation at 94°C for 1 min and annealing /elongation at 66°C for 5 min. Finally, 30 cycles of denaturation at 94°C for 10 s, annealing at 65°C for 50 s and elongation at 72°C for 5 min were performed, with a final hold at 20°C. If necessary, amplification control was performed loading 5 µl of the amplification product on an ethidium bromide stained agarose gel and running electrophoresis for 30 min at 180 V / 80mA. Sanger DNA sequencing itself was run on a 3130xl Genetic Analyzer, using 5 µl Reaction Mix (composed of 0.5 µl BigDye®Terminator v3.1 Cycle Sequencing RR-100, 1.9 µl Q-solution and 2.6 µl water), 5 µl 1:20 diluted amplification product and 5 µl of homemade sequencing primer (exon 1-7; 2.5 pmol/µl). Cycle sequencing assays was performed with a 100-fold approach by a Biomek NXP pipetting roboter. Cycle sequencing itself was run in a SimpliAmp Thermocycler (94°C 2 min, 30 cycles 94°C for 10 s, 60°C 2 min, hold at 20°C). Cycle sequencing reactions were purified using Agencourt CleanSeq magnetic beads in a Biomek NXP protocol following the manufacturer’s instructions. Sanger DNA sequencing itself passed on a 3130xl Genetic Analyzer. After import of sequence raw data in the uType software, the sequences were analyzed for HLA type creation by aligning to recent IMGT HLA allele database. HLA type for asymptomatic seropositive and seronegative donors was determined via surface antibody staining using commercially available antibodies targeting HLA-A*02-FITC (1:200,), HLA-A*03-APC (1:200), HLA-B*07-PE (1:100), HLA-B*08-APCVio770 (1:200).

### 10x genomics for single-cell RNA sequencing

After sorting according to IFN-γ signal via IFN-γ catch (IFN-γ-FITC), cells were centrifuged and the supernatant was carefully removed. Cells were resuspended in the 37,2 µl Mastermix + 37.8 µl water before 70 µl of the cell suspension were transferred to the chip. (Step 1.1 and 1.2 of the original protocol). After each step, the integrity of the pellet was checked under the microscope to ensure that all cells are loaded onto the chip. From here on, 10x experiments have been performed according to the manufacturer’s protocol (Chromium next GEM Single Cell VDJ V1.1, Rev D). Quality control has been performed with a High sensitivity DNA Kit on a Bioanalyzer 2100 as recommended in the protocol and libraries were quantified with the Qubit dsDNA hs assay kit. All steps have been performed using RPT filter tips and DNA LoBind tubes.

For sequencing, libraries have been pooled according to their minimal required read counts (20.000 reads/cell for gene expression libraries and 5.000 reads/cell for TCR libraries). Illumina paired end sequencing was performed with 28+91 bp on a HiSeq2500 or with 2×150 bp on a NovaSeq 6000 for the second experiment. Annotation against the human genome (GRCh38) and a corresponding VDJ reference (vdj_GRCh38_alts_ensembl-3.1.0) was performed using Cell Ranger (V 3.0.2, 10x genomics) for the first experiment. For the second experiment an updated Cell Ranger version (V 5.0.0, 10x genomics) was used in combination with updated references (GRCh38-2020-A and vdj_GRCh38_alts_ensembl-5.0.0)

### Data pre-processing of single-cell RNA sequencing

All detailed information for the strategy of scRNA seq data pre-processing can be found in the uploaded notebooks. All analyses have been performed using SCANPY^68^. Briefly, cells with less than 200 genes were excluded as well as genes present in less than three cells. Cells with more than 20 % mitochondrial genes were excluded and cut-offs for the maximum number of counts (experiment 1: 50.000, experiment 2: 40.000) and number of genes (experiment 1: 7.000, experiment 2: 6.000) were selected individually for the two experiments. Counts were normalized per cell, logarithmized and the variance was scaled to unit variance and zero mean. The number of counts, percentage of mitochondrial genes and cell cycle score was regressed out before highly variable genes were identified and filtered. The data was batch corrected using batch-balanced k nearest neighbors (bbknn) for the individual donors. Donor reallocation was performed using scSplit^41^ and hla-genotyper/gene score for Y chromosome genes (https://www.uniprot.org/docs/humchry.txt) for the first experiment and souporcell^69^ in combination with barcoded antibodies for the second experiment. Since HLA prediction from RNA sequencing data is not very accurate, an HLA score was introduced to allocate donors to clusters derived from scSplit demultiplexing. In principle, HLA matching was scored considering all predictions and all original genotypes in order to find the best match between prediction and genotype. Detailed information can be found in the uploaded notebooks.

### Clonotype definition from single-cell RNA sequencing data

Clonotype analysis was performed using scirpy^70^. Cells belonging to one clonotypes were defined to have identical αand βchain CDR3 nucleotide sequences and both pairs of TRA/TRB sequences were considered in case additional chains have been present.

### TCR DNA template design and CRISPR/Cas9-mediated TCR knock-in

DNA constructs for CRISPR/Cas-9-mediated HDR at TRAC locus were designed *in silico* with the following structure: 5′ homology arm (300–400 base pairs (bp), P2A, TCR-β (including mTRBC with additional cysteine bridge^71^), T2A, TCR-α (including mTRAC with additional cysteine bridge^71^), bGHpA tail, 3′ homology arm (300–400 bp). All HDR DNA template sequences were synthesized by Twist.

CRISPR/Cas9-mediated TCR knock-out and knock-in (KI) with subsequent editing verification via TCR KI staining and FACS was performed as described^42^ on PBMCs isolated from whole blood of healthy donors and cultured at 180 IU/ml IL-2. TCR sequences for re-expression are listed in Supplementary **Fehler! Verweisquelle konnte nicht gefunden werden**..

### pMHC Class I monomer production and biotinylation

All pMHC monomers were produced in-house as previously described^30,72,73^. Briefly, recombinantly expressed and purified human HLA-A*01 and HLA-A*03 heavy chains and β2m were denatured in urea and subsequently refolded into heterodimeric pMHC complexes in presence of an excess of respective synthetic peptide. Re-folded pMHC monomers were purified using size exclusion chromatography, concentrated and stored at -80 °C. pMHC biotinylation was performed as described^30^. Briefly, monomeric pMHCs were activated via a tyrosine tubulin ligase-mediated addition of an azo-tyrosine group and functionalized via click chemistry in presence of DBCO-PEG_4_-Biotin.

### Multimer and TCR KI stainings

For multimer staining, 0.4 µg biotinylated pMHC was multimerized on 0,1 µg fluorophore-conjugated StrepTavidin backbone in a final volume of 50 µl. Up to 5×10^6^ cells were incubated with 50 µl fluorescent-labelled multimers for 45 min on ice protected from light. Surface antibody staining was added after 25 min and incubated for remaining 20 min. Surface marker for TCR KI and multimer stainings covered mTRBC-PE (1:100), CD8-FITC (1:100), CD3-PC7 (1:100).

### RV transduction of A549 HLA editing

A549-ACE2 RFP^+^ cells were kindly provided by the group of Professor Andreas Pichlmair. These cells have been previously engineered to express the SARS-CoV-2 surface receptor protein ACE2 and red fluorescent protein (RFP). A549-ACE2 RFP+ cells were then retrovirally transduced with plasmids encoding HLA-A*01:01 or HLA-A*03:01. Cells were additionally co-transduced with blue fluorescent protein (BFP) for selection of transgene expressing cells. For the production of retroviral particles, RD114 cells were transfected with pMP71 expression vector (containing the HLA heavy chain construct, gag/pol and amphotropic envelope) by calcium phosphate precipitation. Virus supernatant was filtered and added on retronectin-coated-non-tissue treated well plates. 1*10^5^ A549 ACE2 RFP+ cells were then transduced via spinoculation on virus-coated plates. Successfully transduced cells were purity sorted three days after for HLA-A3 expression. In the case of HLA-A1, cells were sorted for BFP expression.

### IncuCyte killing assay

Production and quantification of GFP-expressing SARS-CoV-2 stocks have been performed as previously described74,75. For the Incucyte killing assay, 5*10^3^ A549-ACE2^+^ RFP^+^ HLA-A*01 or A549-ACE2^+^ RFP^+^ HLA-A*03 cells were seeded 24 h prior infection with SARS-CoV-2 GFP virus (MOI 5). Plates were placed in the IncuCyte S3 Live-Cell Analysis System where real-time images of mock (RFP channel) and infected (GFP and RFP channel) cells were captured every 3 h for 72 h. Cell viability (mock and infected), virus growth and infected target cells were respectively assessed as the cell number per image (RFP^+^ objects), total GFP area (µm^2^) per image and overlap of object counts per image (RFP^+^GFP^+^ objects) using IncuCyte S3 Software (Essen Bioscience; version 2019B Rev2). SARS-CoV-2 TCR-engineered T cells were sorted on day 10 post CRISPR/Cas9 editing and were added 24 h post-infection. At endpoint (72 h post-infection), supernatant and T cells were removed, infected cells were washed once with PBS, and fresh medium was added before picture acquisition.

