## Supplementary Data for "Recruitment of highly functional SARS-CoV-2-specific CD8^+^ T cell receptors mediating cytotoxicity of virus-infected target cells in non-severe COVID-19"

**TITLE**

**SUPPLEMENTS**

#### Supplementary Table 1. List of selected SARS-CoV-2derived 9-mer epitopes

| Predicted HLA class I restriction | Peptide name | Sequence | IEDB | Response Frequency<br>Responder/non-responder |
|---|---|---|---|---|
| **HLA-A0101** | **ORF1_DTD** | **DTDFVNEFY** | **1311144** | **7/3 (70%)** |
| **HLA-A0101** | **ORF1_GTD** | **GTDLEGNFY** | **1311156** | **5/5 (50%)** |
| **HLA-A0101** | **ORF1_AMD** | **AMDEFIERY** | **-** | **0/10 (0%)** |
| **HLA-A0101** | **ORF1_LTN** | **LTNIFGTVY** | **-** | **1/9 (10%)** |
| **HLA-A0101** | **ORF3_FTS** | **FTSDYYQLY** | **1309115** | **10/0 (100%)** |
| **HLA-A0101** | **S_LTD** | **LTDEMIAQY** | **1310623** | **5/5 (50%)** |
| HLA-A0101 | S_WTA | WTAGAAAYY | 1327824 | 1/7 (13%) |
| HLA-A0101 | S_CVA | CVADYSVLY* | 7247 | 2/6 (25%) |
| HLA-A0201 | ORF1_KLW | KLWAQCVQL* | 32240 | 10/2 (83%) |
| HLA-A0201 | ORF3_LLY | LLYDANYFL | 1311180 | 8/4 (67%) |
| **HLA-A0201** | **N_LQL** | **LQLPQGTTL*** | **38881** | **0/23 (0%)** |
| **HLA-A0201** | **N_ILL** | **ILLNKHIDA*** | **27182** | **0/23 (0%)** |
| **HLA-A0201** | **N_LLL** | **LLLDRLNQL*** | **37473** | **1/22 (4,3%)** |
| **HLA-A0201** | **S_VLN** | **VLNDILSRL*** | **69657** | **0/23 (0%)** |
| **HLA-A0201** | **S_VVF** | **VVFLHVTYV*** | **71663** | **2/21 (8,6%)** |
| **HLA-A0201** | **S_ALN** | **ALNTLVKQL*** | **2801** | **1/22 (4,3%)** |
| **HLA-A0201** | **S_KVG** | **KVGGNYNYL** | **-** | **0/23 (0%)** |
| **HLA-A0201** | **S_KIA** | **KIADYNYKL** | **1319519** | **7/16 (30%)** |
| HLA-A0201 | S_YLQ | YLQPRTFLL | 1309147 | 11/1 (92%) |
| **HLA-A0301** | **ORF1_KLF** | **KLFDRYFKY** | **1312859** | **5/6 (45%)** |
| **HLA-A0301** | **ORF1_VTN** | **VTNNTFTLK** | **1311232** | **5/6 (45%)** |
| **HLA-A0301** | **ORF1_ASM** | **ASMPTTIAK** | **1310291** | **1/10 (9,1%)** |
| HLA-A0301 | ORF1_KTI | KTIQPRVEK | 1311176 | 6/3 (67%) |
| HLA-A0301 | N_KTF | KTFPPTEPKK* | 33668 | 2/7 (22%) |
| **HLA-A0301** | **S_RLF** | **RLFRKSNLK** | **1075031** | **0/12 (0%)** |
| HLA-A0301 | S_KCY | KCYGVSPTK | 1311170 | 6/3 (67%) |
| HLA-A0301 | S_GVY | GVYFASTEK | 1312627 | 3/6 (33%) |

| | | | | |
|---|---|---|---|---|
| **HLA-A1101** | **ORF1_STF** | **STFNVPMEK** | **1311591** | **2/2 (50%)** |
| **HLA-A1101** | **ORF1_KLF** | **KLFDRYFKY** | **1312859** | **3/1 (75%)** |
| **HLA-A1101** | **ORF1_VTN** | **VTNNTFTLK** | **1311232** | **3/1 (75%)** |
| **HLA-A1101** | **ORF1_ASM** | **ASMPTTIAK** | **1310291** | **3/1 (75%)** |
| **HLA-A1101** | **ORF1_TTI** | **TTIKPVTYK** | **1313808** | **0/4 (0%)** |
| **HLA-A1101** | **M_LSY** | **LSYFIASFR** | **1321267** | **0/4 (0%)** |
| HLA-A1101 | N_ATE | ATEGALNTPK* | 4936 | 3/1 (75%) |
| HLA-A1101 | N_KTF | KTFPPTEPKK | 33668 | 3/1 (75%) |
| HLA-A1101 | S_RLF | RLFRKSNLK | 1075031 | 2/2 (50%) |
| **HLA-A2402** | **ORF1_NYM** | **NYMPYFFTL** | **1310703** | **0/8 (0%)** |
| **HLA-A2402** | **ORF1_YFM** | **YFMRFRRAF** | **1334348** | **0/8 (0%)** |
| **HLA-A2402** | **ORF1_WSW** | **WSMATYYLF** | **1313970** | **0/8 (0%)** |
| HLA-A2402 | ORF1_VYI | VYIGDPAQL* | 72048 | 2/6 (25%) |
| HLA-A2402 | ORF3_VYF | VYFLQSINF | 1310934 | 4/4 (50%) |
| **HLA-A2402** | **M_SYF** | **SYFIASFRL** | **1313718** | **0/8 (0%)** |
| **HLA-A2402** | **N_LSP** | **LSPRWYFYY*** | **39576** | **1/7 (13%)** |
| **HLA-A2402** | **S_PYR** | **PYRVVVLSF*** | **50166** | **0/8 (0%)** |
| **HLA-A2402** | **S_GYQ** | **GYQPYRVVVL*** | **23436** | **0/8 (0%)** |
| **HLA-A2402** | **S_VYS** | **VYSTGSNVF** | **1313944** | **0/8 (0%)** |
| HLA-A2402 | S_QYI | QYIKWPWYI | 1310756 | 5/3 (63%) |
| HLA-A2402 | S_NYN | NYNYLYRLF | 1313269 | 5/3 (63%) |
| HLA-A2402 | S_KWP | KWPWYIWLGF | 1311572 | 1/7 (13%) |
| HLA-A2402 | S_YFP | YFPLQSYGF | 1075121 | 1/7 (13%) |
| **HLA-B0702** | **ORF1_KPN** | **KPNELSRVL** | **1319902** | **1/13 (7,1%)** |
| **HLA-B0702** | **ORF1_KPV** | **KPVETSNSF** | **-** | **0/14 (0%)** |
| **HLA-B0702** | **ORF1_VPM** | **VPMEKLKTL** | **1310914** | **0/14 (0%)** |
| HLA-B0702 | N_SPR | SPRWYFYYL* | 60242 | 11/0 (100%) |
| **HLA-B0801** | **ORF1_FVK** | **FVKHKHAFL** | **1310425** | **1/2 (33%)** |
| **HLA-B0801** | **ORF1_SLS** | **SLSHRFYRL** | **-** | **0/3 (0%)** |
| **HLA-B0801** | **ORF1_YLK** | **YLKLRSDVL** | **-** | **0/3 (0%)** |
| HLA-B1501 | N_AQF | AQFAPSASA* | 3956 | 5/3 (63%) |
| **HLA-B3501** | **ORF1_VPF** | **VPFWITIAY** | **1326965** | **1/1 (50%)** |
| **HLA-B3501** | **ORF1_FAV** | **FAVDAAKAY** | **1331939** | **0/2 (0%)** |
| **HLA-B3501** | **ORF1_LVA** | **LVAEWFLAY** | **1321432** | **1/1 (50%)** |
| **HLA-B3501** | **ORF1_NVL** | **NVLEGSVAY** | **-** | **0/2 (0%)** |
| **HLA-B3501** | **ORF1_HSI** | **HSIGFDYVY** | **-** | **1/1 (50%)** |
| HLA-B3501 | S_QPT | QPTESIVRF | 1323461 | 1/1 (50%) |

| | | | | |
|---|---|---|---|---|
| HLA-B3501 | S_LPF | LPFNDGVYF | 1321049 | 2/0 (100%) |
| HLA-B4010 | M_SEL | SELVIGAVIL | 1075044 | 2/0 (100%) |
| HLA-B4010 | N_MEV | MEVTPSGTWL* | 190494 | 2/0 (100%) |
| HLA-B4410 | N_MEV | MEVTPSGTWL* | 190494 | 6/3 (67%) |
| HLA-B4410 | ORF1_SEF | SEFSSLPSY* | 57432 | 2/7 (22%) |
| HLA-C0702 | N_QRN | QRNAPRITF | 1309136 | 1/12 (7,7%) |

*100% homology with SARS-CoV-1

**Bold print peptides were part of initial 9-mer peptide pool.**

**Supplementary Table 2. Clinical information study cohorts**

| Study cohort | Gender | Age | SARS-COV-2 PCR+ |
|---|---|---|---|
| Mild symptomatic (N=53) | Female: 32 (60%)<br>Male: 19 (36%)<br>Unknown: 2 (4%) | 40,5 ± 10,6 | yes |
| Seropositive (N=28) | Female: 18 (64%)<br>Male: 10 (36%) | 38,4± 13,5 | no |
| Seronegative (N=37) | Female: 23 (62%)<br>Male: 14 (38%) | 38,8± 13,6 | no |
| Unexposed pre-pandemic (N=28) | Female: 12 (55%)<br>Male: 10 (45%) | 42,96±12,9 | no |

**Supplementary Table 3. SARS-CoV-2 TCR sequences**

| TCR | IFN-γ cluster | TRAV | TRAJ | TRA_1_cdr3 | TRBV | TRBJ | TRB_1_cdr3 |
|---|---|---|---|---|---|---|---|
| 13 | positive | TRAV13-1 | TRAJ47 | CAAFGNKLVF | TRBV16 | TRBJ1-5 | CASSHSNSNQPQHF |
| 18 | negative | TRAV21 | TRAJ56 | CAVDTGANSKLTF | TRBV12-3 | TRBJ2-5 | CASSLMTIQETQYF |
| 28 | positive | TRAV12-1 | TRAJ31 | CVVRNNNARLMF | TRBV9 | TRBJ2-1 | CASSVDGSSYNEQFF |
| 32 | positive | TRAV12-2 | TRAJ9 | CAPLGGRRGFKTIF | TRBV7-8 | TRBJ2-1 | CASSGGTSGSHNEQFF |
| 43 | positive | TRAV12-2 | TRAJ17 | CAVSGGRAAGNKLTF | TRBV7-9 | TRBJ2-6 | CASSAGSGANVLTF |
| 82 | negative | TRAV8-2 | TRAJ12 | CVVSRMDSSYKLIF | TRBV13 | TRBJ2-2 | CASSSSGGQTLTGELFF |
| 3398 | positive | TRAV20 | TRAJ35 | CAVQAEGFGNVLHC | TRBV2 | TRBJ2-2 | CASSEPTSGELFF |
| 3399 | negative | TRAV36/DV7 | TRAJ42 | CAVETYGGSQGNLIF | TRBV13 | TRBJ1-1 | CASSAQGAGTEAFF |
| 3409 | negative | TRAV5 | TRAJ5 | CAENRTGRRALTF | TRBV20-1 | TRBJ1-1 | CSGEAALNTEAFF |
| 3456 | positive | TRAV38-2/DV8 | TRAJ38 | CAYIYAGNNRKLIW | TRBV7-6 | TRBJ2-1 | CASSSDGGGFNEQFF |

#### SUPPLEMENTARY FIGURE LEGENDS

**Supplementary Fig. 1. *Ex vivo* SARS-CoV-2 specific T cell responses are at detection limit in convalescent donors.** PBMCs were isolated from mild COVID-19 individuals 4-6 weeks after infection. 5 x $10^6$ PBMCs were stimulated with 1 µg/µl Peptivator S pool (15-mer) or in-house designed 9-mer peptide pool for 4 h and stained for IFN-γ cytokine release. Representative raw data (A) and quantification (B) of SARS-CoV-2-specific T cells (IFN-$\gamma^+$) are depicted. IFN-γ-reacting cells were pre-gated on $CD3^+$. Responder were identified respective to the non-stimulated negative control and background subtraction.

**Supplementary Fig. 2. Detection of SARS-CoV-2 specific $CD8^+$ T cells after i*n vitro* expansion in mild COVID-19 individuals.** 5 x $10^6$ PBMCs from mild COVID-19 individuals were stimulated with 9-mer peptide pool-pulsed autologous PBMCs (10 µg/µl) and expanded for 12 days prior to re-challenge (1 µg/µl). Exemplary raw data (A) and quantification (B) of IFN-γ release after 4 h re-stimulation with 9-mer peptide pool in $CD8^+$ and $CD8^-$ T cells at day 0 and day 12 of expansion. Statistical analyses were performed via paired-parametric test (** p value < 0.01).

**Supplementary Fig. 3. Comparison of $CD8^+$ and $CD8^-$ T cell responses after stimulation with SARS-CoV-2 9-mers.** 5 x $10^6$ PBMCs from four indicated cohorts were stimulated with 9-mer peptide pool-pulsed autologous PBMCs (10 µg/µl) and expanded for 10-12 days prior to re-challenge (1 µg/µl). IFN-γ release of CD8+ and CD8- cells, pre-gated on CD3+ living lymphocytes, was assessed after 4 h re-stimulation. Statistical analyses were performed via paired-unparametric Wilcoxon test (* p value < 0.05, ** p value < 0.01, *** p value < 0.001, **** p value < 0.0001).

**Supplementary Fig. 4. Frequency of HLA class I molecules among the study cohorts.** **A)** Frequency of HLA class I molecules in cohort of mild COVID-19 (red) and pre-pandemic healthy individuals (grey) determined via next generation sequencing. **B)** Frequency of HLA class I molecules in cohort of asymptomatic seropositive (blue) and seronegative (orange). assessed via antibody staining, due to limited sample accessibility and according to commercial availability.

**Supplementary Fig. 5. $CD8^+$ T cell responses to SARS-CoV-2 peptide subpools.** 5 x $10^6$ PBMCs from mild COVID-19 individuals were stimulated with 9-mer pool-pulsed autologous PBMCs (10 µg/µl) and expanded for 10-12 days. Expanded PBMCs were re-stimulated with 1 µg/µl peptide sub-pools (each containing 8-10 peptides) for 4 h and reactivity was measured as percentage of IFN-γ-releasing $CD8^+$ T cells. Heat map shows the percentage of $CD8^+$ T cells producing IFN-γ in response to peptide subpool stimulation (red gradient scale).

**Supplementary Fig. 6. IFN-γ responses to individual peptide stimulation.** 5 x $10^6$ PBMCs from mild COVID-19 individuals were stimulated with 9-mer pool-pulsed autologous PBMCs (10 µg/µl) and expanded for 10-12 days. Expanded PBMCs were re-stimulated with 1 µg/µl individual peptides for 4 h and stained for IFN-γ release. Depicted are raw data showing reactivity of $CD8^+$ and $CD8^-$ T cells to the 19 immunogenic SARS-CoV-2 epitopes of the initial in-house designed peptide pool from different donors. For flow-cytometry analyses, reactive cells were pre-gated on $CD3^+$ living lymphocytes.

**Supplementary Fig 7. Response rate to SARS-CoV-2 $CD8^+$ T cell epitopes according to HLA class I genotype A)** Bar graph showing the percentage of individuals with detectable SARS-CoV-2-specific $CD8^+$ T cells according to HLA class I molecules. HLA-coverage of initial peptide selection (top graph; Table 1, bold print) and HLA-coverage after inclusion of recently published SARS-CoV-2 immunogenic epitopes (bottom graph). Responses to individual SARS-CoV-2 epitopes predicted for the same HLA class I molecule were pooled. **B)** Heat map showing the percentage of $CD8^+$ T cells producing IFN-γ in response to individual peptide stimulation (orange-black gradient scale) and the number of reactive epitopes per donor (blue gradient scale) for peptides of the follow-up selection and recently published epitopes (Table 1, regular print) in mild COVID-19 cohort. Responder and non-responder were defined according to IFN-γ response to initial peptide selection (Table 1, bold print).

**Supplementary Fig. 8. Gating strategy for flow cytometry-based cell sorting for scRNA-seq.** After 12 days *in vitro* expansion on SARS-CoV-2 9-mer peptide pool, PBMCs from mild COVID-19 individuals were re-stimulated with 1µg/µl crude A1/ORF1_VTN or A3/ORF3a_FTS epitopes for 4 h. Cells were stained for CD3, CD8 and IFN-γ and sorted on an MoFlo Astrios EQ . For each donor, 2.500 $CD8^+$IFN-$\gamma^+$ and 10.000 $CD8^+$IFN-$\gamma^-$ T cells were sorted. HLA-A*01:01 donors 32 and 22, and HLA-A*03:01 donors 22 and 34 were sorted in the same well prior processing for scRNA-seq.

**Supplementary Fig. 9. Strategy for sample demultiplexing and donor assignment. A**) Principal component analyses on single-nucleotide-polymorphisms (SNPs) colored by the clusters identified with scSplit. **B)** Donor identification from clusters according to Y chromosome genes. Cluster 0_0 was defined as male gender due to enrichment in the male gene score. **C)** Donor identification from clusters according to HLA annotation. HLA predictions from hla-genotyper for each cluster were compared to the original HLA type of the pooled donors to generate the HLA score. **D)** UMAPs showing the distribution of cells of the UMAP for the individual donors. The gray background represents the total dataset, the color corresponds to the Leiden clustering. **E)** UMAPs depict the individual donors. Highlighted are the re-expressed TCRs from each donor.

**Supplementary Fig. 10. Functional assessment for IFN-γ$^-$ cluster-derived TCRs.** PBMCs from the same donors the transgenic TCRs were isolated from were pulsed with 10 µg/µl SARS-CoV-2 9-mer pool for 2 h and co-cultured with TCR-engineered T cells for 4 h, prior intracellular staining of IFN-γ and IL-2.

**Supplementary Fig. 11. Identification of signatures for peptide reactivity and functionality of antigen-specific TCRs. A)** Volcano plot shows the differentially expressed genes between reactive and non-reactive TCRs to SARS-CoV-2 epitopes. Selected genes for the reactivity score are colored black. **B)** Results for linear regression analysis between $EC_{50}$ values of functional TCRs and single genes. Genes selected for the functionality score are colored black. Dashed lines represent the selection criteria for genes to be included in the functionality score. **C)** Linear regression for the individual genes selected from (B) for all TCRs (upper panel) and only functional TCRs (lower panel). The gray Area in (C) depicts the 95% confidence interval. * p value < 0.05, ** p value < 0.01, *** p value < 0.001, **** p value < 0.0001. Two-sided p-values for a hypothesis test (null hypothesis: slope is zero) have been calculated in (j-l), using Wald Test with t-distribution of the test statistic.

**Supplementary Fig. 12. Demultiplexing of samples according to HTO-antibodies.** UMAPs were generated according to HTO counts for each cell. The heatmap corresponds to Leiden clustering or counts of one individual HTO-antibody.

#### SUPPLEMENTARY MATERIALS AND METHODS

#### KEY RESOURCES TABLE

| REAGENT or RESOURCE | SOURCE | IDENTIFIER |
|---|---|---|
| **Antibodies for flow cytometry** | | |
| CD19-ECD (J3-119) | Beckman Coulter | A07770 |
| CD8-PE (3B5) | Life Technologies | MHCD0804 |
| CD8-eF450 (OKT8) | Life Technologies | 48-0086-42 |
| CD8-FITC (B9.11) | Beckman Coulter | A07756 |
| CD3-BV421 (SK7) | BD Biosciences | 56797 |
| CD3-PC7 (UCHT.1) | Beckman Coulter | 737657 |
| mTRBC-PE (h57-597) | Biolegend | 109208 |
| mTRBC-APC/Fire750 (H57-597) | Biolegend | 109246 |
| IFN-γ FITC (25723.11) | BD Biosciences | 340449 |
| HLA-A*02-FITC | BD Biosciences | 551285 |
| HLA-B*07-PE | Biolegend | 372404 |
| HLA-A*03-APC | Miltenyi | 130-115-795 |
| HLA-B*08-APCVio770 | Miltenyi | 130-099-5910 |
| Streptavidin-APC | eBioscience | 17-4317-82 |
| Total Seq-C 0251 (10x Hashtaq 1) | Biolegend | 394661 |
| Total Seq-C 0252 (10x Hashtaq 2) | Biolegend | 394663 |
| Total Seq-C 0253 (10x Hashtaq 3) | Biolegend | 394665 |
| Total Seq-C 0254 (10x Hashtaq 4) | Biolegend | 394667 |
| Total Seq-C 0255 (10x Hashtaq 5) | Biolegend | 394669 |
| Total Seq-C 0256 (10x Hashtaq 6) | Biolegend | 394671 |
| IFN-γ catch | Miltenyi | 130-090-433 |
| **Cell Culture Reagents** | | |
| RPMI 1640 Gibco | Sigma | R0883 |
| DMEM | Life Technologies | 10938025 |
| Pancoll human (1.077g/ml) | PAN Biotech | P04-601000 |
| Fetal calf serum | Biochrom | |
| Human serum | In house | |
| β-mercaptoethanol | Life Technologies | 31350010 |
| Gentamicin | Life Technologies | 15750-037 |
| HEPES | Life Technologies | 15630056 |
| L-Glutamine | Sigma | G8540-100G |
| Penicillin/Streptomycin | Life Technologies | 10378016 |
| Recombinant human IL-2 | Peprotech | 200-02 |
| Recombinant human IL-7 | Peprotech | 200-07 |
| Recombinant human IL-15 | Peprotech | 200-15 |
| Phorbol myristate acetate | Sigma | P1585 |
| Ionomycin | Sigma | I9657 |
| Fibronectin | Sigma | F2006 |
| **Chemicals, Peptides, Enzymes, Kits** | | |
| Propidium Iodide (PI) | Life Technologies | P1304MP |
| Ethidium Monoazide Bromide (EMA) | Life Technologies | E1374 |
| Cytofix/Cytoperm | BD Biosciences | 554714 |
| GolgiPlug | BD Biosciences | 555029 |
| DMSO | Merck | D8418 |
| DBCO-PEG4-Biotin | Jena Bioscience | CLK-A105P4-10 |
| Retronectin | TaKaRa | T100B |
| BigDye®Terminator v3.1 Cycle Sequencing RR-100 | Applied Biosystems | |
| Q-solution | Qiagen | |
| Agencourt CleanSeq magnetic beads | Beckman Coulter | |
| Alt-R® S.p. HiFi Cas9 Nuclease V3 | IDT | 1081061 |
| Alt-R® Cas9 Electroporation Enhancer | IDT | 1075916 |
| Alt-R® CRISPR-Cas9 crRNA | IDT | |

| | | |
|---|---|---|
| 5′-AGAGTCTCTCAGCTGGTACA-3′ for TRAC | | |
| Alt-R® CRISPR-Cas9 crRNA 5′-GGAGAATGACGAGTGGACCC-3′ for TRBC | IDT | |
| Alt-R® CRISPR-Cas9 tracrRNA | IDT | 1072532 |
| Agencout AMPure XP | Beckman Coulter | A63881 |
| High sensitivity DNA Kit | Agilent | 5067-4626 |
| Qubit dsDNA hs assay kit | Life Technologies | Q32851 |
| RPT filter tips | Starlab | S1183-1710, SS1180-8710, S1182-1730 |
| DNA LoBind tubes | Sigma | EP0030108051, EP0030108078, EP0030124359 |
| P3 Primary Cell Kit | Lonza | V4XP-3024<br>V4SP-3096 |
| PepTivator SARS-CoV-2 Protein S | Miltenyi | 130-126-701 |
| SARS-CoV-2 individual peptides | IBA<br>peptides & elephants | |

| Reference sequences | NCBI ID |
|---|---|
| SARS-COV-2 (Wuhan-Hu-1) | NC_045512 |
| SARS-CoV | NC_004718 |
| MERS-CoV | NC_019843 |
| HCoV-OC43 | NC_006213 |
| HCoV-HKU1 | NC_006577 |
| HCoV-NL63 | NC_005831 |
| HCoV-229E | NC_002645 |

| Serology | Source | Identifier |
|---|---|---|
| iFlash SARS-CoV-2 IgG (2019-nCov IgG) | SHENZHEN YHLO BIOTECH CO | C86095G |

| Experimental Models: Cell Lines & Virus | |
|---|---|
| RD114 | In house |
| K562-HLA-A*01:01 BFP | In house production |
| K562-HLA-A*03:01 BFP | In house production |
| A549-ACE2-RFP-HLA-A*01:01 | In house production |
| A549-ACE2-RFP-HLA-A*03:01 | In house production |
| SARS-CoV-2 GFP | In house production |

| Hardware | |
|---|---|
| Äkta pureSuperdeso 200 10/300GL | GE |
| SimpliAmp Thermocycler | Applied Biosystems, Darmstadt, Germany |
| 3130xl Genetic Analyzer | Applied Biosystems, Darmstadt, Germany |
| Biomek NXP pipetting roboter | |
| 4D-Nucleofector | Lonza |
| CytoFlex S Cell Analyzer | Beckman Coulter |
| MoFlo Astrios EQ | Beckman Coulter |
| 2100 Bioanalyzer | Agilent |
| NovaSeq 6000 | Illumina |
| Incucyte S3 Live-Cell Analysis System | Sartorius |

| Software and Algorithms | |
|---|---|
| NETMHC4.0 | NetMHC 4.0 Server (dtu.dk) |

| | |
|---|---|
| Netstab1.0 | NetMHCstab 1.0 Server (dtu.dk) |
| NETCTL1.2 | NetCTL 1.2 Server (dtu.dk) |
| PickPocket1.1 | PickPocket 1.1 Server (dtu.dk) |
| NetMHCpan4.1 | NetMHCpan 4.1 Server (dtu.dk) |
| IEDB T cell epitope prediction tools | T Cell Tools (iedb.org) |
| FloJo V10 | FlowJo LLC |
| GraphPad Prism 9 | Graphpad |
| Microsoft Excel | Microsoft |
| Affinity Designer 1.9 | Serif |
| IncuCyte S3 Software, Version 2019B Rev2 | Essen, Bioscience |
| uType software | Invitrogen/ThermoFisher |
| Scanpy 1.4.3 | Wolf et al (https://doi.org/10.1186/s13059-017-1382-0) |
| Scirpy 0.3 | Sturm et al. (https://doi.org/10.1093/bioinformatics/btaa611) |
| scSplit 1.0 | Xu et al, https://doi.org/10.1186/s13059-019-1852-7 |
| Souporcell 2.0 | Heaton et al (https://doi.org/10.1038/s41592-020-0820-1) |
| Hla-genotyper 0.4 | https://pypi.org/project/hla-genotyper/ |
| Cell Ranger 3.0.2/5.0.0 | https://support.10xgenomics.com/single-cell-gene-expression/software/pipelines/latest/installation |

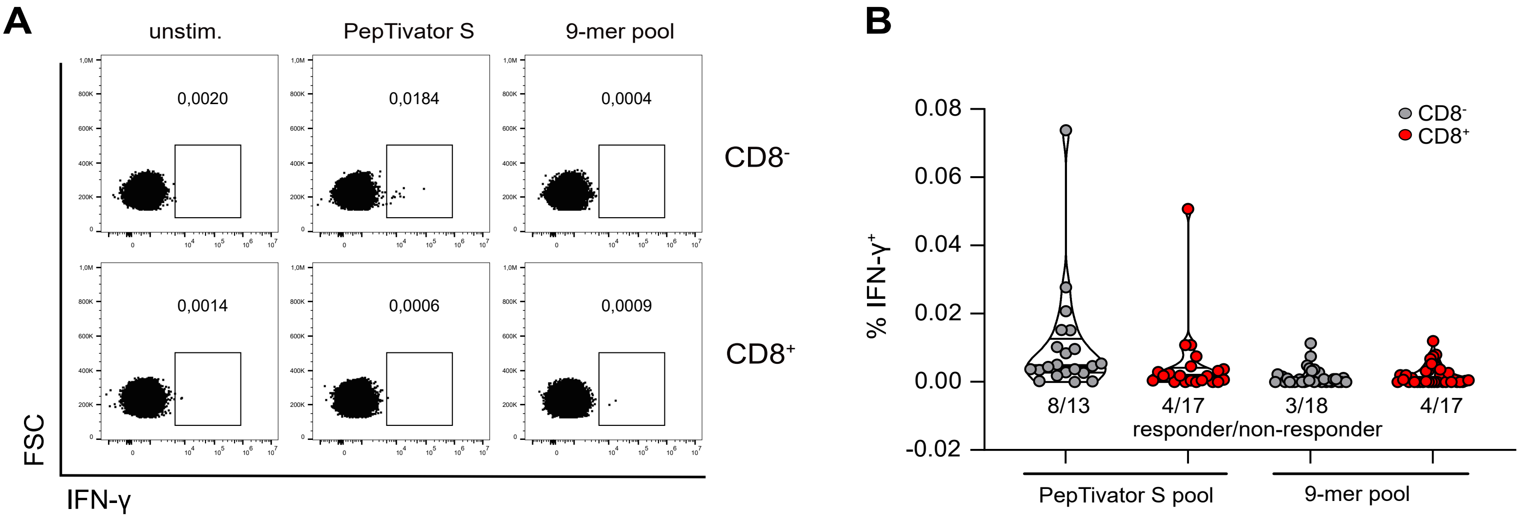


**Supplementary Fig. 1**

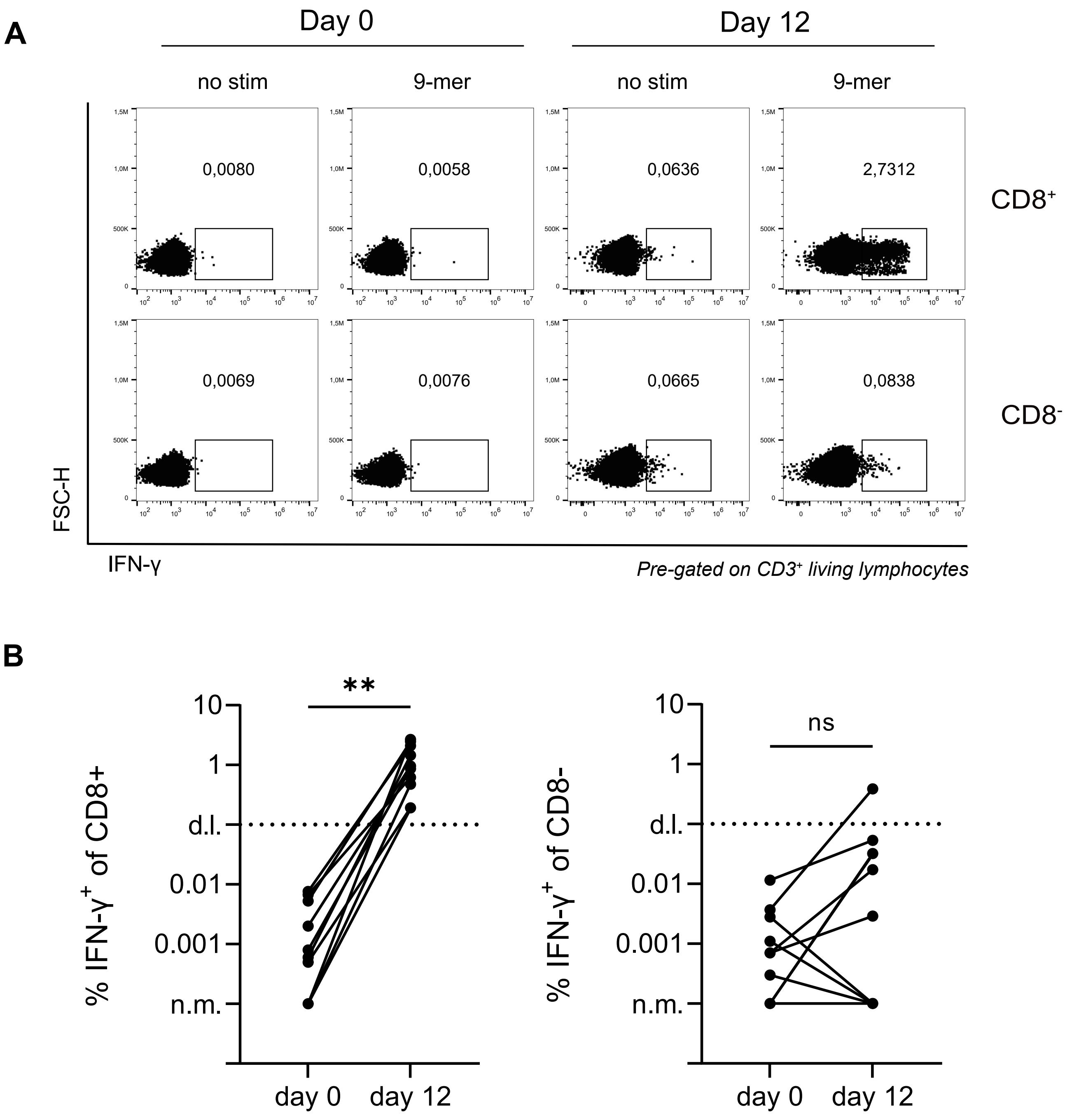


**Supplementary Fig. 2**

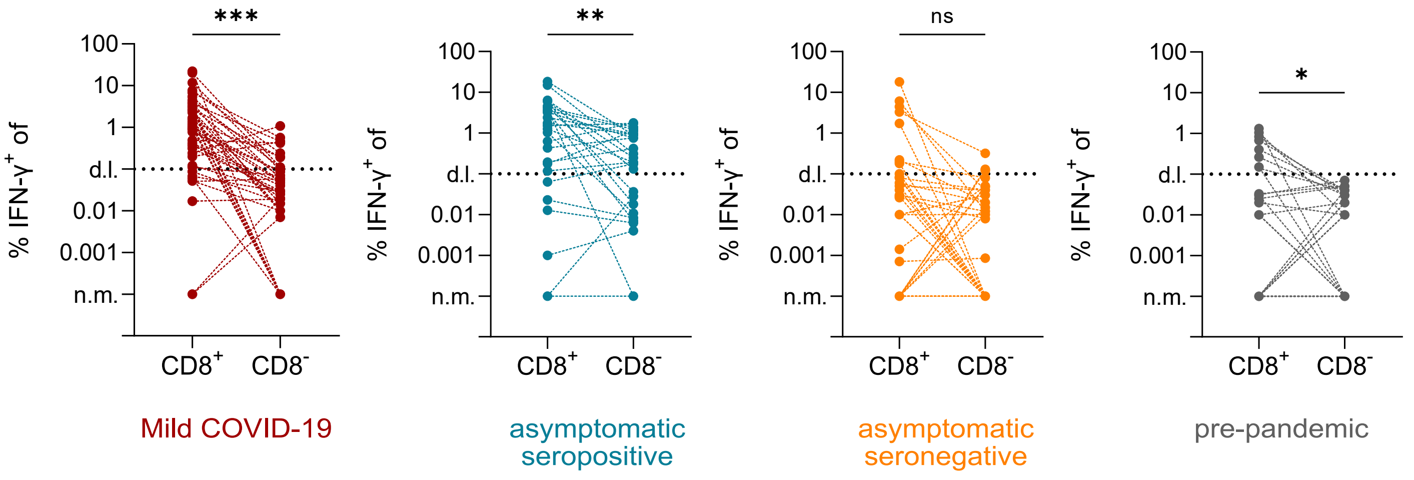


#### Supplementary Fig. 3

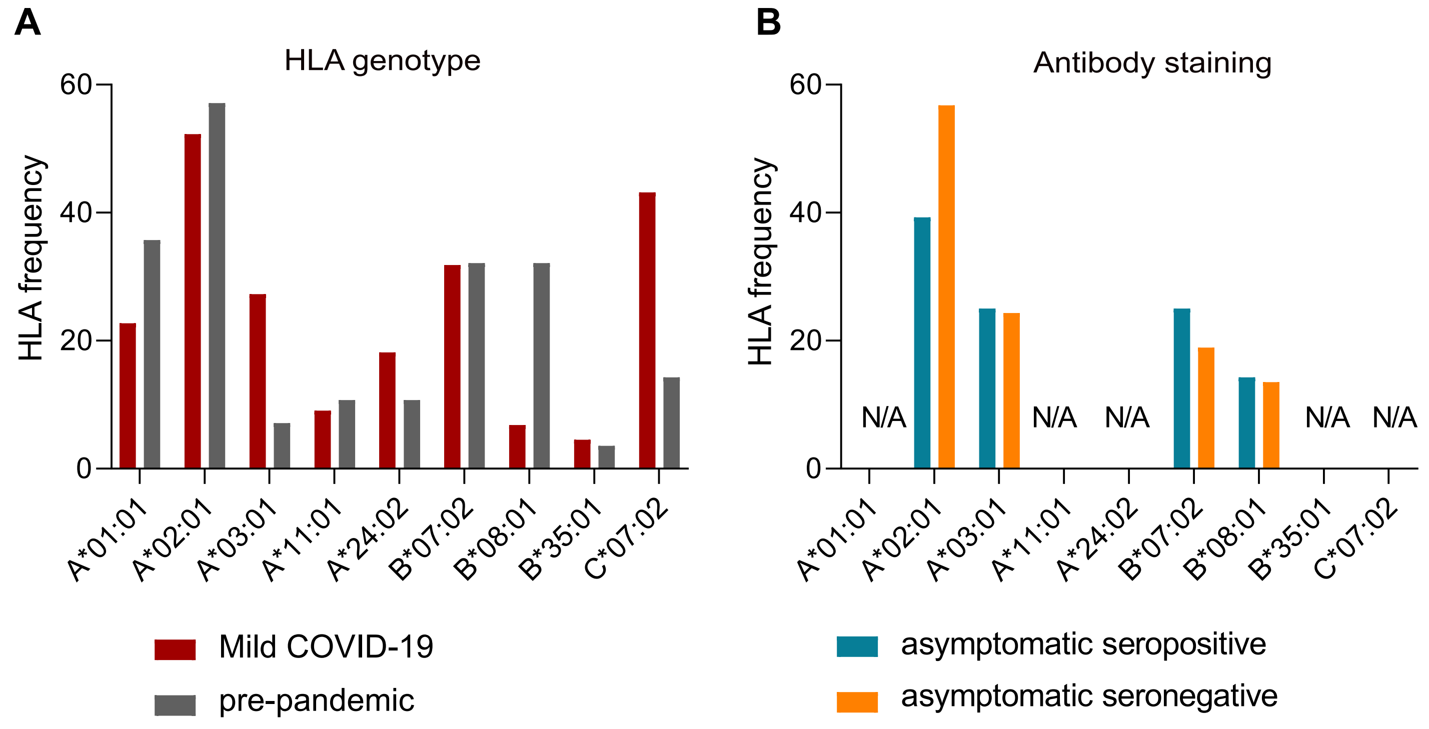


**Supplementary Fig. 4**

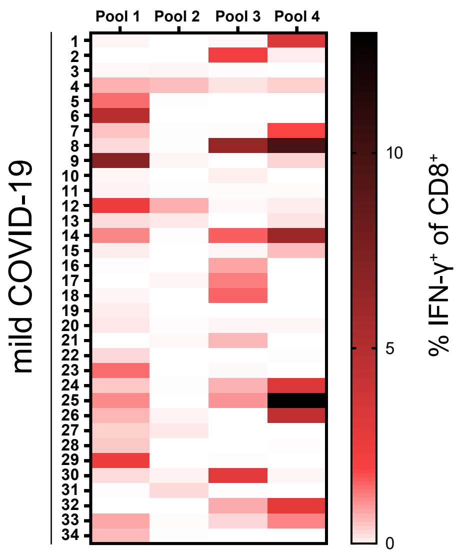


**Supplementary Fig. 5**

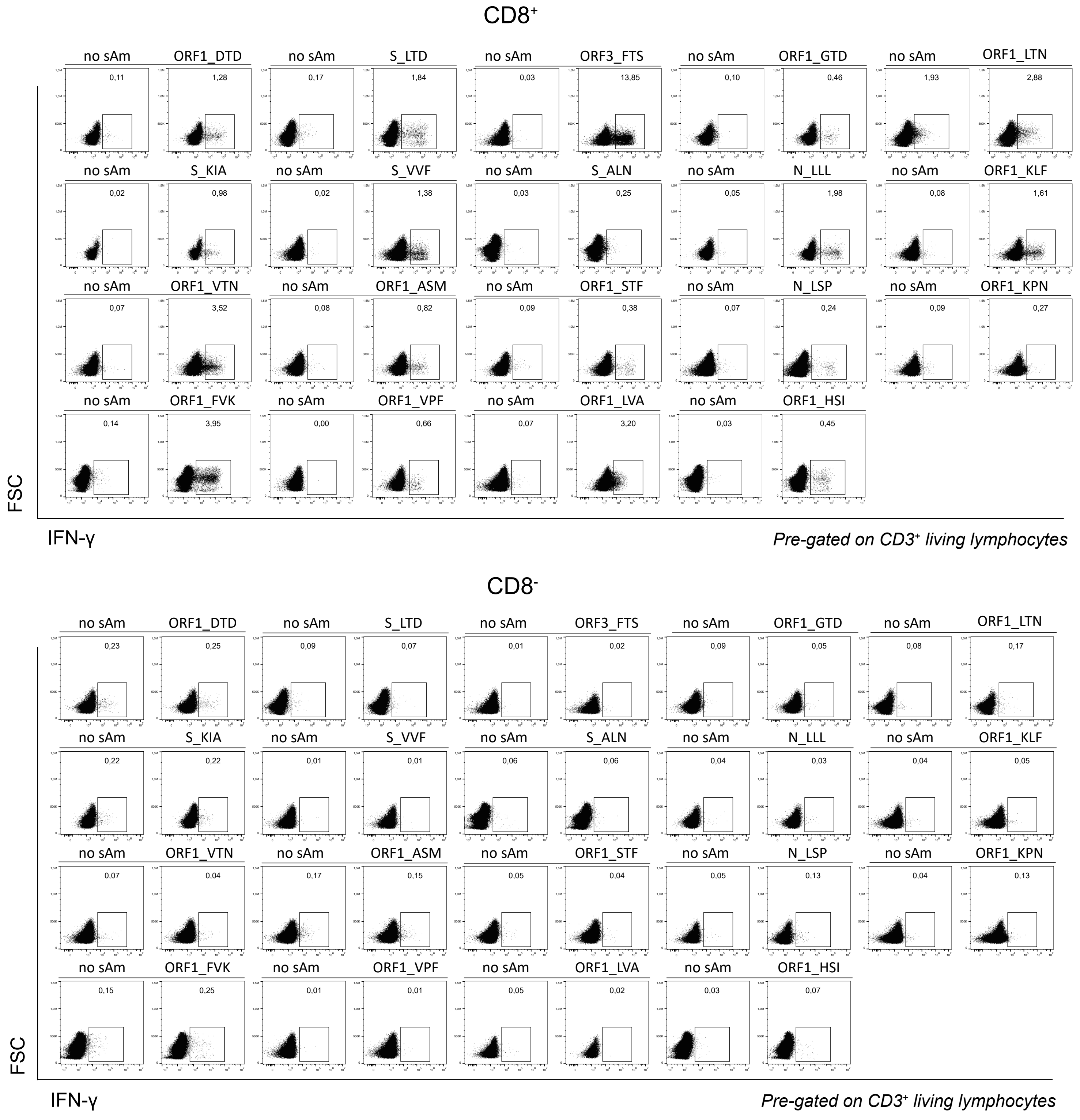


**Supplementary Fig. 6**

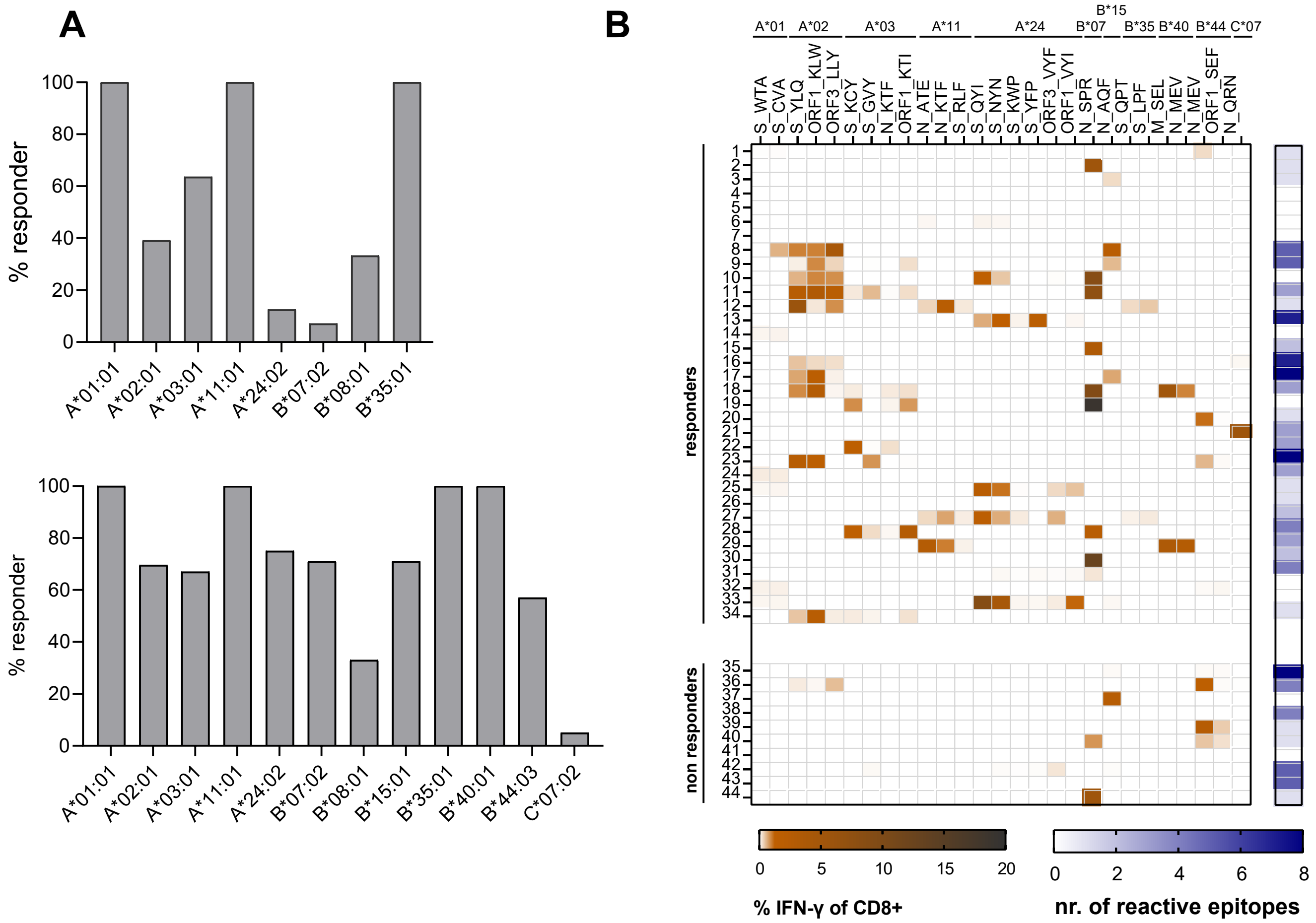


**Supplementary Fig 7**

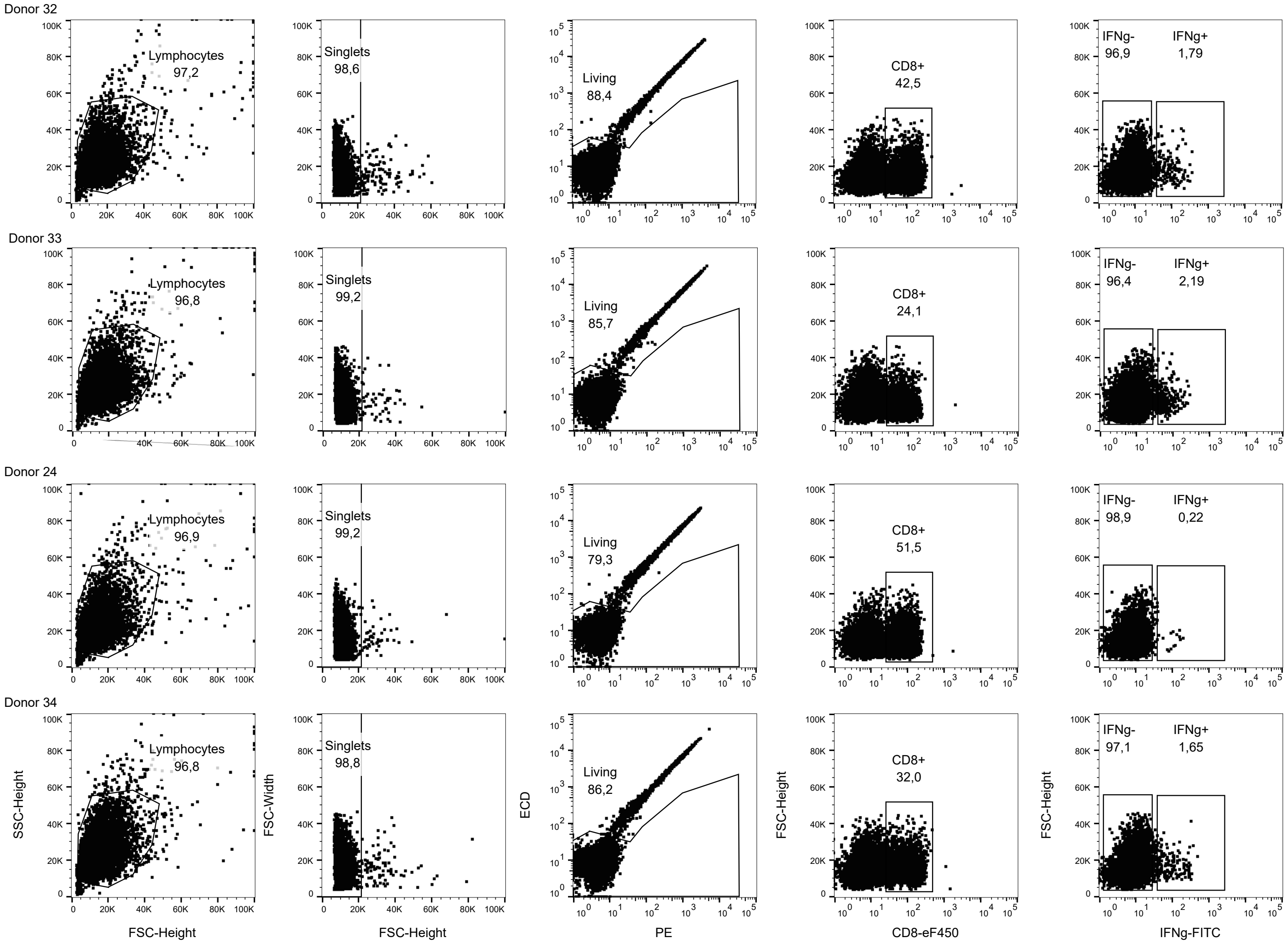


### Supplementary Fig. 8

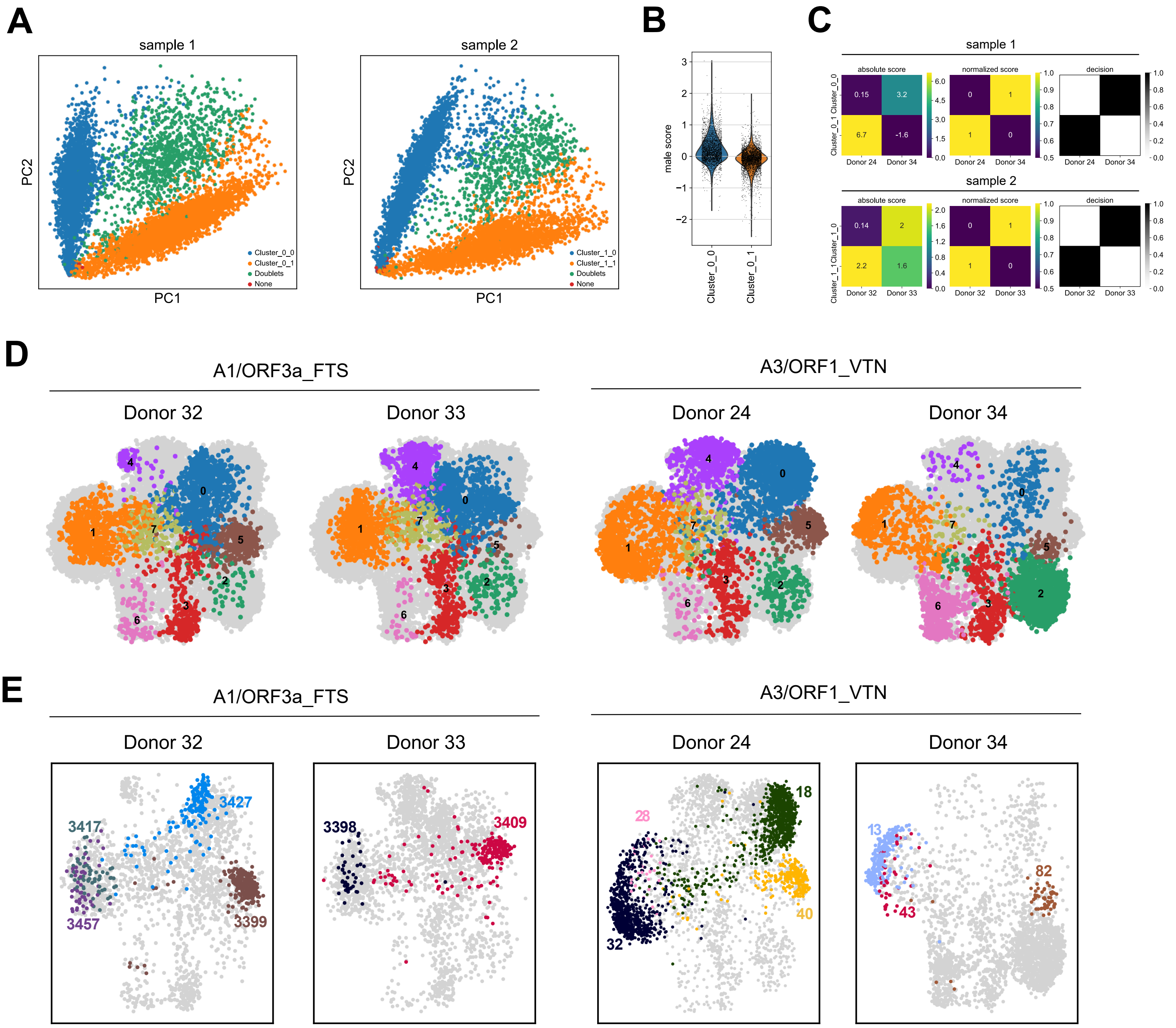


**Supplementary Fig. 9**

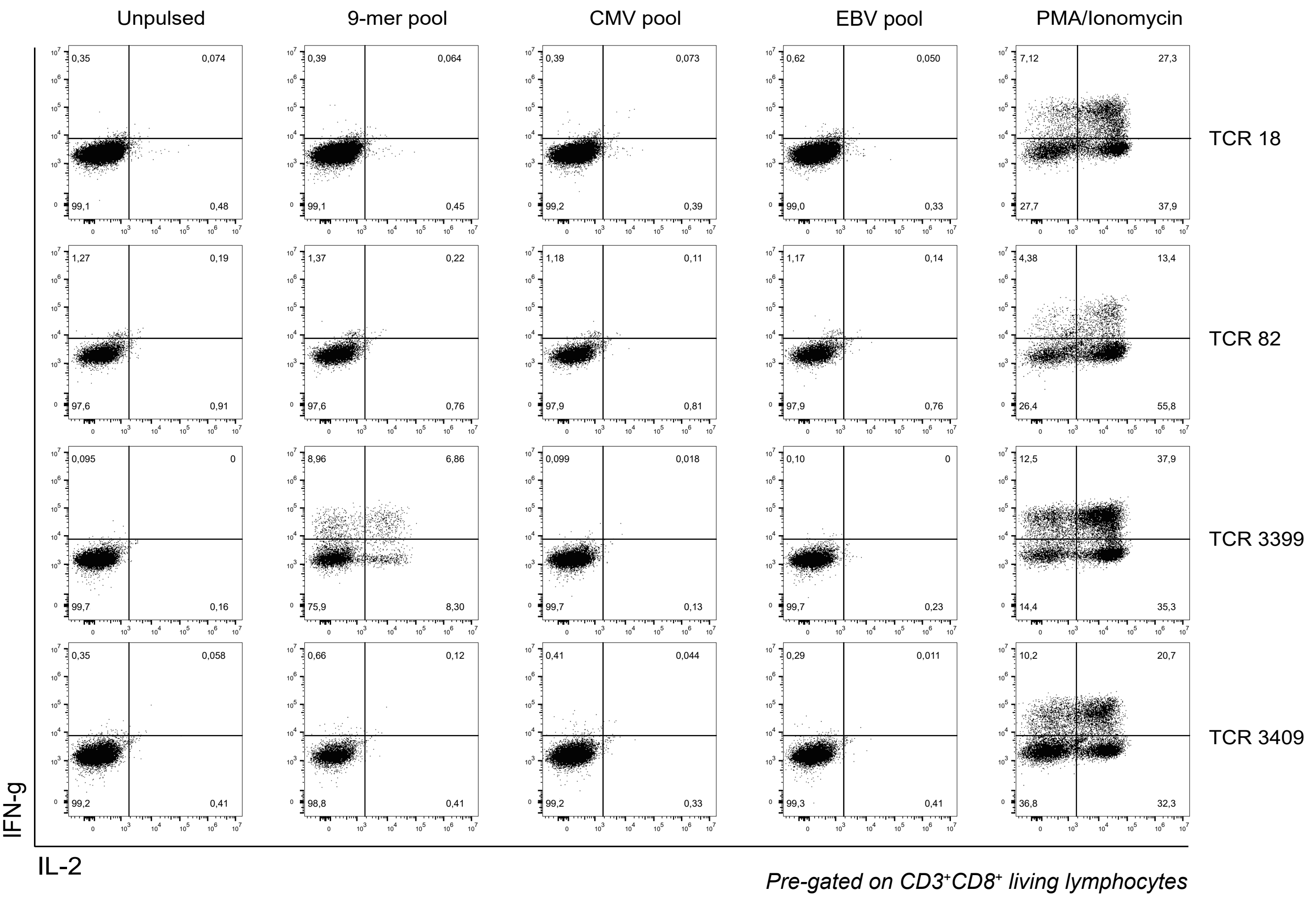


**Supplementary Fig. 10**

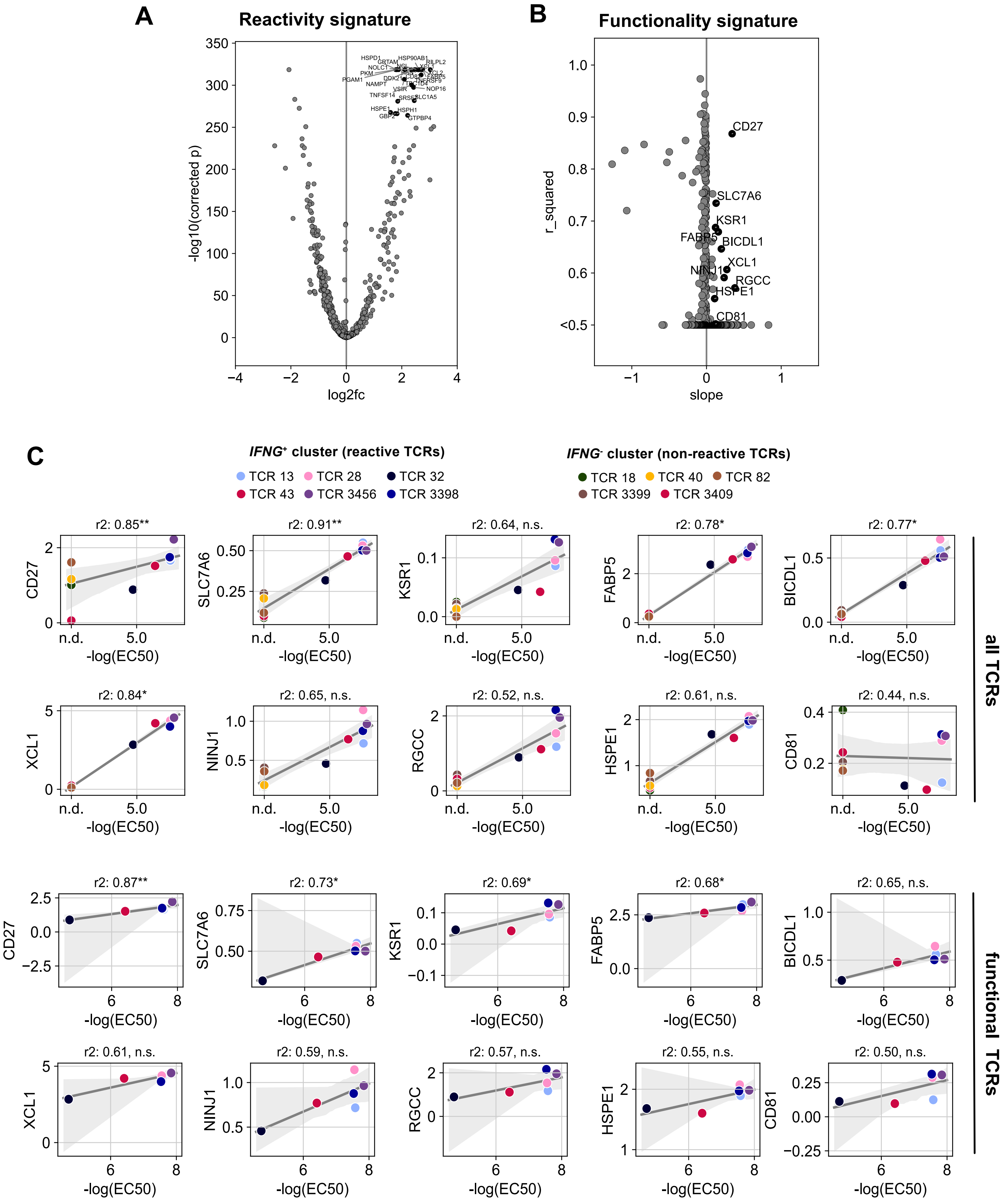


**Supplementary Fig. 11**

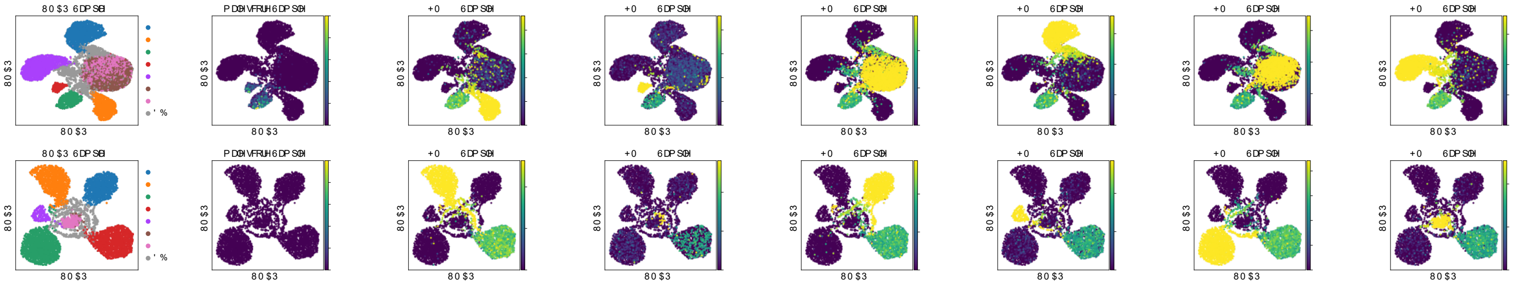

### Supplementary Fig. 12
